# Using Explainable-AI to Find Geospatial Environmental and Sociodemographic Predictors of Suicide Attempts

**DOI:** 10.1101/2022.04.26.22274333

**Authors:** Mirko Pavicic, Angelica M. Walker, Kyle A. Sullivan, John Lagergren, Ashley Cliff, Jonathon Romero, Jared Streich, Michael R. Garvin, MVP Suicide Exemplar Workgroup, the Million Veteran Program, John Pestian, Benjamin McMahon, David W. Oslin, Jean C. Beckham, Nathan A. Kimbrel, Daniel A. Jacobson

**Affiliations:** Oak Ridge National Laboratory, Computational and Predictive Biology, Oak Ridge, TN; The Bredesen Center for Interdisciplinary Research and Graduate Education, University of Tennessee Knoxville, Knoxville, TN; Cincinnati Children’s Hospital Medical Center, University of Cincinnati, Cincinnati, OH; Theoretical Biology and Biophysics, Los Alamos National Laboratory, Los Alamos, NM; VISN 4 Mental Illness Research, Education, and Clinical Center, Center of Excellence, Corporal Michael J. Crescenz VA Medical Center, Philadelphia, PA, USA; Department of Psychiatry, Perelman School of Medicine, University of Pennsylvania, PA, USA; Durham Veterans Affairs Health Care System, Durham, NC; VA Mid-Atlantic Mental Illness, Research, Education, and Clinical Center; Department of Psychiatry and Behavioral Sciences, Duke University Medical Center, Durham, NC, USA; Duke University School of Medicine, Duke University, Durham, NC; VA Health Services Research and Development Center of Innovation to Accelerate Discovery and Practice Transformation, Durham, NC

**Author notes:** Contributed equally (Mirko Pavicic, Angelica M. Walker).

## Abstract

Despite a global decrease in suicide rates in recent years, death by suicide has increased in the United States. It is therefore imperative to identify the risk factors associated with suicide attempts in order to combat this growing epidemic. In this study, we use an explainable-artificial intelligence method, iterative Random Forest, to predict suicide attempts using data from the Million Veteran Program. Our predictive model incorporates multiple environmental variables (e.g., elevation, light wavelength absorbance, temperature, humidity, etc) at ZIP code-level geospatial resolution. We additionally consider demographic variables from the American Community Survey as well as the number of firearms and alcohol vendors per 10,000 people in order to assess the contributions of proximal environment, access to means, and restraint decrease to suicide attempts. Our results show that geographic areas with higher concentrations of married males living with spouses are predictive of lower rates of suicide attempts, whereas geographic areas where males are more likely to live alone and to rent housing are predictive of higher rates of suicide attempts. We also identified climatic features that were associated with suicide attempt risk by age group. Additionally, we observed that firearms and alcohol vendors were associated with increased risk for suicide attempts irrespective of the age group examined, but that their effects were small in comparison to the top features. Taken together, our findings highlight the importance of social determinants and environmental factors in understanding suicide risk among veterans.

## Introduction

Suicide rates in the United States (U.S.) have been increasing in recent years despite these rates declining globally^1^. According to the biopsychosocial model of suicide risks, there are distal, developmental, and proximal factors that can affect the probability of suicide attempt^2^. Distal factors are those related to familial and genetic predisposition and early-life adversity. Developmental factors include personality traits associated with suicidal behavior, cognitive deficits, and chronic substance misuse. Proximal, or precipitating factors include but are not limited to psychiatric, psychological, socioeconomic, and environmental factors. Even though socioeconomic and environmental factors are classified as proximal factors, they can simultaneously influence distal and developmental factors such as early-life adversity and individual development. Several studies have found associations between a number of demographic factors and suicide such as age, ethnicity, socioeconomic status, marital status, religion, etc.^3^. Furthermore, climatic variables such as cumulative solar radiation has been negatively associated with suicide risk, while temperature and diurnal temperature range have been positively associated with suicide risk^4–6^. However, to thoroughly analyze the effect of environmental and socioeconomic factors on an individual-level risk of attempting suicide, a comprehensive study at high geospatial and temporal resolution is needed to account for within-county variation^7^.

Other relevant proximal factors include access to means and substance misuse^8,9^. For example, occupations with access to lethal means are associated with increased risk of death by suicide and method of choice^10,11^. Moreover, controlling access to lethal means is an effective strategy for decreasing suicide risk^12^. In the U.S., death by suicide is the leading cause of violent deaths, and firearms are responsible for approximately half of these deaths^13^. Substance misuse also plays an important role in suicide prevention because acute substance intoxication can increase an individual’s disinhibition. For example, a prior study showed that suicide decedents have an increased risk of alcohol ingestion and intoxication before their death relative to controls^14^.

The objective of the present research was to conduct an analysis of climatic and socio-demographic features that are associated with increased risk for suicide attempts among U.S. veterans using an explainable-artificial intelligence (X-AI) model. We were also interested in the relationship of the number of firearms and alcohol vendors per 10,000 people as proxies for access to means and decreased restraint, which were also included in the final model. Together, we identify a number of novel factors at zip code-level resolution that impact individual-level risk for attempting suicide.

## Methods

### Data and Data Pre-processing

#### Patient Data

The cohort used in this study was the same as previously described^15^, with some modifications. The patients in this study were enrolled in the U.S. Department of Veterans Affairs’ (VA) Million Veteran Program (MVP). All subjects provided informed consent and the research presented here were approved by the VA Central Institutional Review Board. Cases were defined as having a history of one or more suicide attempts (including both fatal and non-fatal), whereas controls were defined has having no history of suicide attempts or ideation. This vector was created from electronic health records (EHR) from the VA corporate data warehouse (CDW), using International Classification of Diseases (ICD) diagnostic codes, survey data from the mental health domain, and the Suicide Prevention Application Network (SPAN) data set^16^. We encoded these data as a binary vector, where controls were represented by 0 and suicide attempts were set to 1, regardless of whether the attempt was or was not fatal. In total, this resulted in 405,540 patients with 391,409 controls and 14,131 attempts. Each attempt and control were associated with climatic and socio-demographic features according to the most recent patient ZIP code. Sex distribution was 8% female and 92% male. This cohort was 73% white, 18% black or African American, 1% American Indian or Alaska Native, 1% Asian, and 7% mixed race, missing, or unknown. The mean age was 62.4 years for the whole cohort, 63.4 for males and 50.8 years for females (**Supplementary Figure 1A**). Notably, this cohort was unbalanced in terms of cases and controls where only 3% attempted suicide versus 97% who did not, which is reflective of the occurrence of suicide attempts in the general population. Most of the suicide attempts in this cohort were concentrated around 60 years of age, likely since this age group is overrepresented among these patients (**Supplementary Figure 1A and B**). However, the proportion of attempts grouped by age decreased steadily after age 60 (**Supplementary Figure 1C**). Due to this rapid decrease in attempts after age 60, we explored the socio-demographic and climatic features that were associated with suicide attempts in patients greater than 60 years of age separately from those under 60 years of age. The split with patients above or equal to 60 contained a total of 267,447 individuals with 4,231 attempts and 263,216 controls. The split with patients below age 60 was composed of 138,093 individuals with 9,900 suicide attempts and 128,193 controls.

#### Climatic Features

The climatic features included two groups: static measurements and monthly measurements. Monthly measurements included longitudinal features such as monthly average precipitation and maximum temperature. There were 12 distinct measurement types recorded each month, totaling 144 features. The 30 static features included features such as elevation and percent urban cover. This led to a total of 174 climatic and weather-related features, mapped to 33,144 ZIP codes across the U.S.^17–23^ (**Supplementary Table 1**).

#### Socio-demographic Features

The socio-demographic features were collected from the 2019 American Community Survey, produced by the United States Census Bureau^24^. These features included demographic, socioeconomic, and housing features, such as the percentage of the population that were white females between the age of 30 and 34 living in the ZIP code, percentage of rented housing units in the ZIP code that have adequate plumbing facilities, and the Gini Index of Income Inequality. These 1606 features were captured using the tidycensus software package in R using 5 years estimate for 2019^25^. These features were normalized to represent a percentage of the total population or age bracket within each ZIP code. We also included two additional features: population density (people per square mile) and the ratio of water to land area, which led to a total of 1608 demographic features that were mapped to 33,120 ZIP codes across the U.S (**Supplementary Table 2**).

#### Alcohol and Firearms Features

The Historical Business Database^26^ contains information on the businesses that exist in every ZIP code in the U.S. We utilized two important features from this dataset: one that measured the number of firearms vendors per 10,000 residents in each ZIP code, averaged across the years 2010 and 2019, and the similarly calculated number of alcohol vendors. This information was included for 31,378 ZIP codes across the U.S (**Supplementary Table 3**).

### Explainable-AI Analysis

In this study, we used iterative Random Forest (iRF)^27,28^, an X-AI algorithm that ranks input features by importance through iterative feature weighting, to associate proximal environmental features with suicide attempts. Thus, data pre-processing was necessary to ensure accuracy and objectivity in the results by reducing the number of highly correlated, redundant features explaining variance in the model. Large pairwise correlations between input features may negatively impact the iRF model, since the importance of one feature is then split across the correlated features, thereby decreasing their overall importance in predicting suicide attempt. To account for this, if one feature was highly correlated with another, one of the features was selected to be a representative of both features, while the other feature was removed from consideration. Pearson’s correlation coefficients were calculated between all 1,784 features. Pairs of features with a correlation coefficient greater than or equal to 0.90 in absolute value were considered to be highly correlated. This analysis found 390 features that were highly correlated with each other. Of these 390 features, 142 of them were kept as representative features (**Supplementary Table 4**). Thus, 248 features were dropped, reducing the total number of features to 1,536.

### iRF k-Fold Cross Validation and Accuracy Calculation

To obtain accuracy scores for the iRF model, k-fold cross validation was used. For this study, 5-fold cross validation was repeated ten times, for a total of 50 computational runs. For each run, the data was separated into an 80/20 training/testing split, where 80% of the data was used for training the model and the remaining 20% was reserved for testing. To prevent geographical bias, patients within the same ZIP code were moved to either the training or testing dataset. Prediction accuracy was calculated using the average Area Under the Precision Recall curve (AUPRC), where precision was defined as *true positives / true positives + false positives* and recall as *true positives / true positives + false negatives*. Each random forest in the iRF model includes 1000 trees with a leaf node size of 1000 patients. We set the number of iRF iterations to five to rank the importance of each feature in predicting suicide attempt. In addition to ranking input features by importance, we next identified the feature-level explainability of our model by determining whether each feature was predictive for or against suicide attempt. To estimate if a feature predicts suicide attempt or controls, the result of each split was averaged using the given feature and mapping those to a linear effect. This provided both the feature effect in the slope of the line and an R^2^ with how closely that related to each of the set of splits. If the slope of the line was positive, then the feature effect size direction was positive, i.e., the value and the feature were positively correlated. If the slope of the line was negative, then the feature effect size direction was negative.

### Model selection

Initially we ran iRF models with *k*-fold cross validation using all 1,536 features to capture the most important features predicting suicide attempt in patients above or equal 60 years and below 60 years of age. Then, we used several combinations of the most important features together with alcohol and firearm vendors per 10,000 residents to run reduced iRF models with *k*-fold cross validation to evaluate if this improved the accuracy of the models. The accuracy of the reduced models was better than using all features in all cases as shown in the area under the precision recall curves (AUPRC) for all 50 data splits used to test the models (**Supplementary Figure 2 and 3**). These results show that by using all features we may introduce noise that may reduce accuracy and explainability of the model. Interestingly, most of the models had some degree of predictability, since most of them had AUPRC above random chance, and this effect was stronger in the age group of patients below 60 years of age (red dashed line in **Supplementary Figure 2**). Based on these results, we selected the model including the top 20 features plus firearms and alcohol vendors for model explanation.

### iRF-LOOP

To show how features or groups of features interconnect each other, we applied iRF-Leave One Out Prediction (iRF-LOOP), which is an extension of the iRF model^27^. In this framework, iRF was used to compute all-against-all predictions of each vector of data from all other vectors. The results of this analysis were captured as networks, in which nodes (i.e., features) were connected by an edge if the pair of features were predictive of each other, thereby revealing functional relationships and subgroups within and across data layers. We performed iRF-LOOP using the pre-processed input matrix which consisted of climate, census, and alcohol and firearm business data, with a total of 1,536 features and 31,378 samples or ZIP codes. This analysis created an all-to-all directed feature association network that captured the relationships between data layers. The resulting network was then thresholded to the top 1% of edges to capture the most important connections between features.

## Results

### Features that were most strongly associated with suicide attempts

iRF predictive models can compute the importance of each feature predicting the dependent variable as well as the direction of the prediction, which in this case was predicting suicide attempt vs control. We examined iRF models trained with 1,536 features to identify the 20 most important predictive features in patients below, and above or equal to age 60, plus *firearms and alcohol vendors per 10,000 residents* (**Supplementary Figure 4 and 5**). The resulting features were further analyzed to determine directionality (i.e., if they predict suicide attempts or controls) and can be aggregated in groups related to emotional support, housing, ancestry, commuting and mobility, access to healthcare, cognitive difficulties, access to means, decrease restraint and climate (**Figure 1 and 2**).

**Figure 1.**
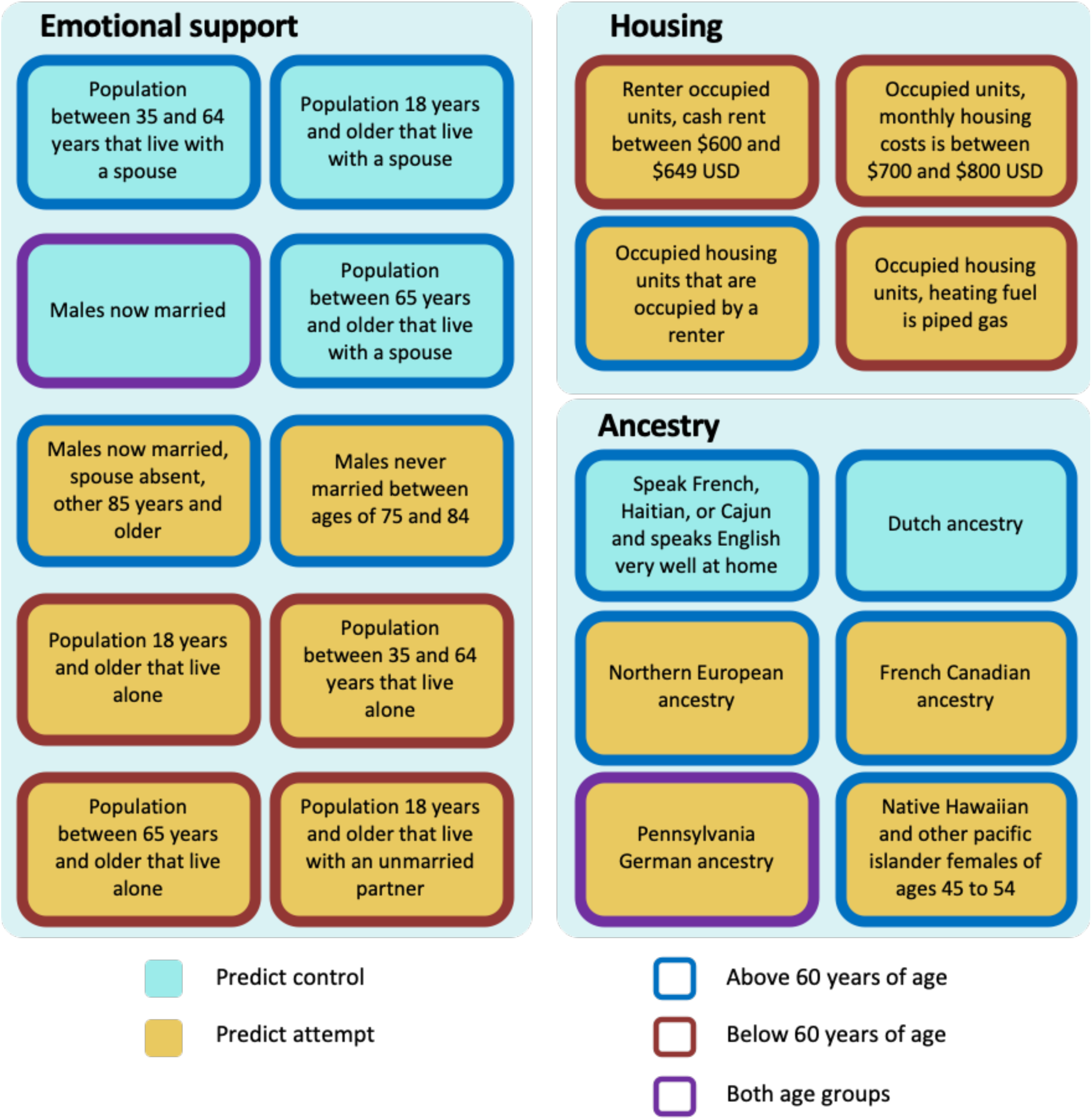
Summary of features found by iRF related to emotional support, housing, and ancestry. Cyan: predict control, yellow: predict attempt, blue border: equal to/above 60 years of age, red border: below 60 years of age, purple border: both age groups.

**Figure 2.**
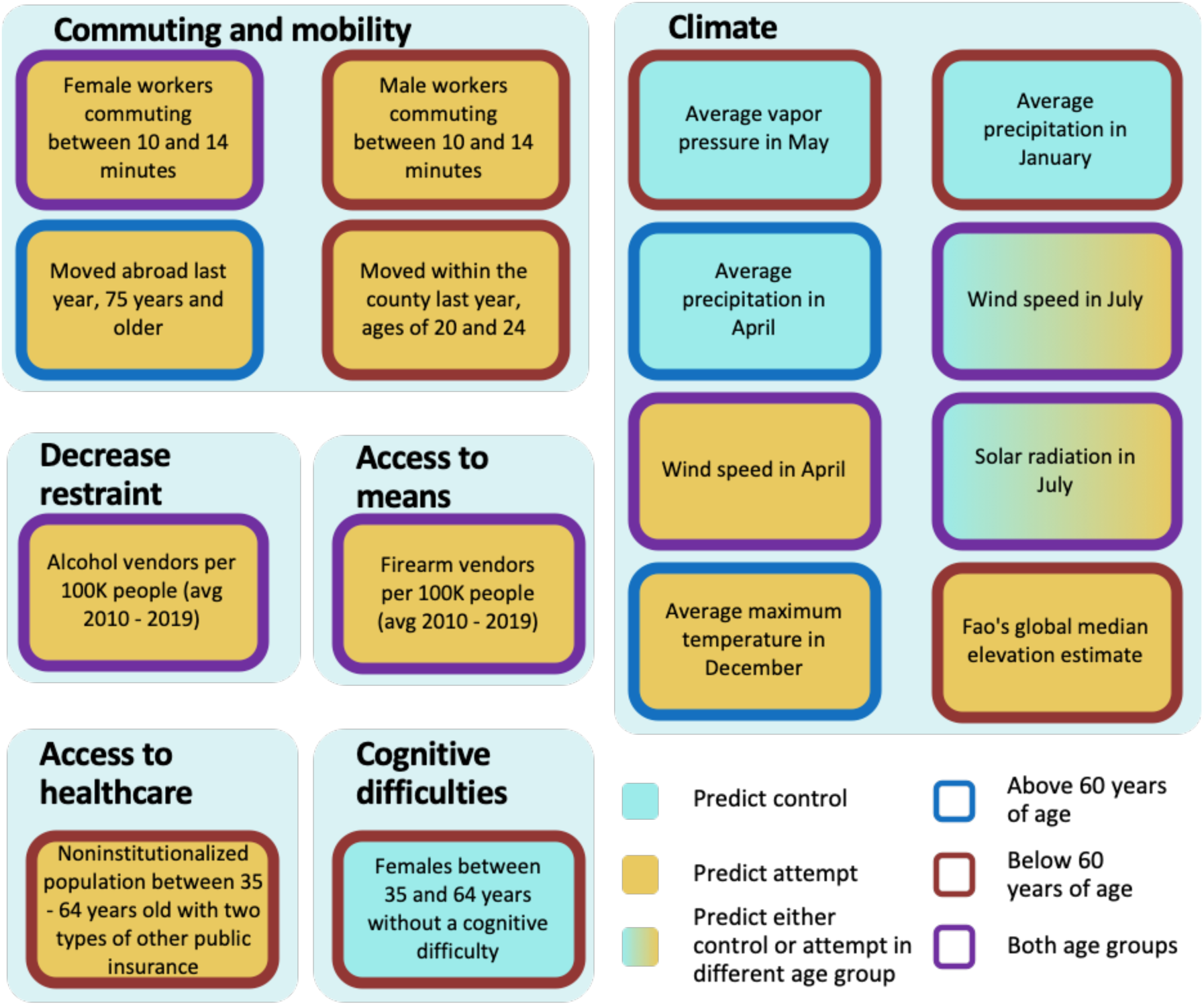
Summary of features found by iRF related to commuting and mobility, climate, decreased restraint, access to means, access to healthcare and cognitive difficulties (proxy for disabilities). Cyan: predict control, yellow: predict attempt, blue border: equal to/above 60 years of age, red border: below 60 years of age, purple border: both age groups.

In the model using patients of above or equal 60 years of age, the features *married males* and *household with spouse present between 35 - 64, and 65 years of age and above* were associated with decreased risk for attempting suicide, whereas *males never married of 75 - 84 years*, and *married males of 85 years of with spouse absent* were associated with increased risk for attempting suicide (**Figure 1**). Ancestry also appeared among the top features explaining suicide attempts. *French Canadian, Northern European*, and *Pennsylvania German ancestries* were associated with increased risk for suicide attempts, whereas *Dutch ancestry* and *speaking French (Haitian or Cajun dialects) at home* were associated with reduced risk (**Figure 1**). *Females with a work commute lasting 10 to 14 minutes*, and *number of firearms* and *alcohol vendors per 10,000 residents* were also associated with increased risk (**Figure 2**). Several climate features were also predictive: *precipitation in April, solar radiation in July (which represents solar radiation in* June, through September), and *wind speed in July* (which represents wind speed in May through September) were associated with decreased risk, whereas *wind speed in April* (which represents wind speed for the rest of the year) and *maximum temperature in December* (which represents average temperature from October through April, minimum temperature from November through March, and maximum temperature from October through March) were associated with increased risk **(Figure 2**). For representative relationships among variables (Pearson > 0.9) see Supplementary Table 4.

For the model using only individuals under 60 years of age, the two most important features associated with decreased risk were *married males*, and *females with no cognitive difficulties between 35 and 64 years of age* (which represents a lack of disabilities, including no hearing, vision, ambulatory, or self-care difficulties in females 35-64 years of age) (**Figure 1 and 2**). Conversely, the features *living alone* and *living with unmarried partner* were associated with increased risk (**Figure 1**). Similarly, *commuting between 10 to 14 mins*, regardless of gender, *monthly housing costs between $700-800, cash rent between $600-649, house heating using gas, moved within county*, and *having two types of public insurance* were all associated with increased risk (**Figure 1 and 2**). On the other hand, only *Pennsylvania German ancestry* was associated with increased risk in the ancestry category (**Figure 1**). Similarly to the model from patients 60 years of age or above, *number of firearm* and *alcohol vendors per 10,000 inhabitants* were associated with increased risk (**Figure 2**). Regarding climatic features, *precipitation in January* (which represents precipitation December through February) and *water vapor in May* (which represents water vapor from April through October and minimum temperature in May) were associated with reduced risk (**Figure 2**). Contrary to the first model, in patients under 60 years of age the climatic variables *solar radiation in July (which represents solar radiation in* June through September), and both *wind speed in July* (which represents wind speed in May through September) and wind speed in *April* (which represents wind speed during the rest of the year) were predictive of individuals with a history of suicide attempt (**Figures 1 and 2**). Additionally, *terrain elevation* were also associated with suicide attempts (**Figure 2**).

### Feature network by iRF-LOOP

It is well known that alcohol abuse increases suicide risk^2^. Thus, we included the number of alcohol vendors per 10,000 people in our final model. Interestingly, when used in combination with the top 20 features, alcohol vendors per 10,000 people ranked 12th and 20th in feature importance in patients above or equal to 60 and below 60 years of age respectively, even though it ranked 360th (above age of 60) and 74th (below age of 60) in the models using all features (**Supplementary Figure 4 and 5**). This could mean that other features are competing with number of alcohol vendors per 10,000 people within the iRF model to classify suicide attempt and controls. Therefore, we created a feature network using iRF-LOOP to explore how features relate to each other. **Figure 3** shows the immediate neighbors of the feature measuring the number of alcohol vendors per 10,000 people, where each node in the subnetwork represents a feature and the arrows represent the edges (connections) between features. Edges are weighted by their normalized feature importance value and the arrow direction shows what feature is predicting another one. Several features related to geographic mobility, population density, low gross rent with cash, and high home value were observed. Further, the subnetwork shows associations among features including number of widowed females, several European ancestries, and the population-adjusted density of alcohol vendors. Regarding climate features, minimum temperature in July and maximum temperature in December were observed.

**Figure 3.**
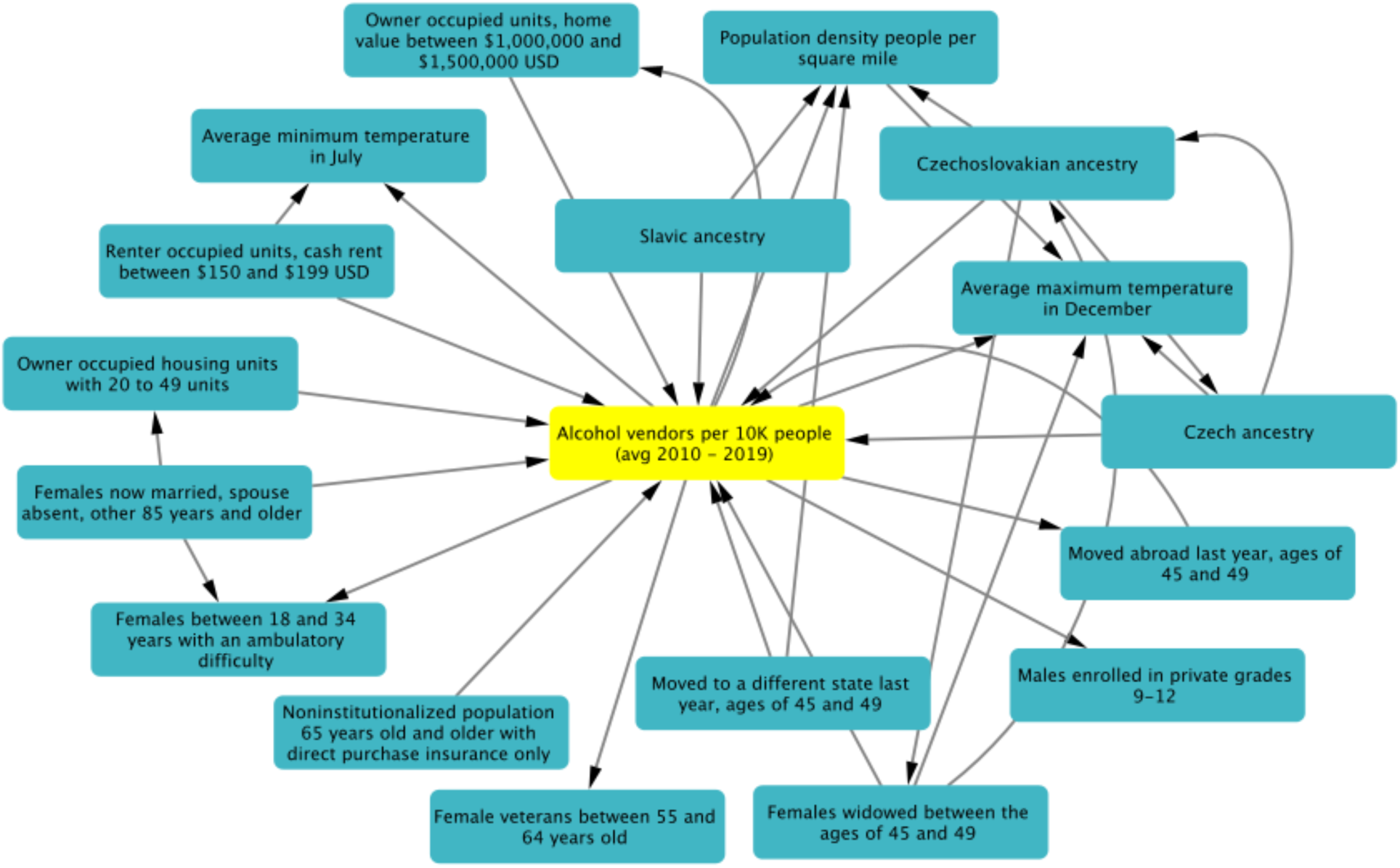
iRF-LOOP Subnetwork showing first neighbors of alcohol vendors per 10000 people. Only the top 1% of edges weighed by normalized importance are shown. Arrow direction corresponds to one feature predicting another feature.

## Discussion

Most prior suicide prevention studies have focused on a relatively small number of features. Moreover, most have typically relied on individual-level information only (e.g., clinical features) or aggregated data for a given geographical area with low geospatial resolution. In the present study we showed that ZIP code-level data can improve prediction in individuals with a history of suicide attempt greater than random chance, supporting the role of the surrounding environment as proximal/precipitating factors that influence the propensity for an individual to attempt suicide. Furthermore, using X-AI we were able to screen more than a one thousand seven hundred demographic and climatic features to obtain several that appear to potentially protect or increase veterans’ propensity for attempting suicide.

An individual’s environment can have profound effects on their psychological state and subsequent risk of suicide. In fact, it has been well documented that socioeconomic and demographic factors such as poverty, education and population density are related to suicide risk^29–31^. In our study we used the EHR data from VA CDW, ACS and climate data to build a predictive model of suicide attempts in the U.S. veteran population. It is worth highlighting that ACS and climate data are not direct information from the individuals under study, but rather a representation of the environment in which they currently reside. Moreover, in the cohort used here, there was a rapid decline in the number of suicide attempts after the age of 60 (**Supplementary Figure 1**), which is in agreement with a surveillance summary from the Center for Disease Control and Prevention (CDC), where the highest suicide rates were observed in age group between 35 and 64 years^32^. Thus, we divided the population into cohorts below or equal to/above 60 years of age for further analysis, since age groups may be affected differently by risk factors^33^.

Interestingly, the top features can be classified into nine main groups namely: emotional support, housing, ancestry, commuting and mobility, climate, decreased restraint, access to means, cognitive difficulties and access to healthcare (**Figure 1 and 2**). In the emotional support group, the protective effect of marital status on suicide risk is well-documented^34^. Studies have shown that married people showed the lowest rate of suicide rates^34–36^, whereas divorced or separated persons are twice as likely to commit suicide than married persons, and this effect is stronger in divorced males^37^. In our study, for all age groups we observed that living in areas with high proportions of married males and individuals living with a spouse were protective factors against suicide attempts for those 60 years of age or above (similar to the individual-level protective factor of marital status). Conversely, areas with high proportions of individuals living alone and unmarried males were associated with higher rates of suicide attempts^38^.

Regarding the commuting and mobility group, commuting time has been associated with depression in a dose responsive manner^39^. Our results showed that living in areas with high proportions of individuals with commutes between 10 and 14 minutes has been associated with increased risk for suicide attempts, irrespective of age. The iRF-LOOP network showed that commuting features are predictive of each other regardless of the commuting time (**Supplementary figure 6**). Thus, commuting between 10 and 14 minutes may be acting as a proxy for commuting in general. We also observed housing-related features for both age groups in relation to suicide attempts. These results are consistent with Lorant *et al*. (2005)^40^, who showed that higher education and home ownership status decreases the risk of suicide. Other studies have found that living in rented units increased the risk for suicide in middle-aged males and females^31^. This effect might be especially important for females who live in large urban areas^41^.

In the ancestry group, we observed that living in areas with a higher proportion of Northern European ancestry was associated with suicide attempts. Although suicide attempts vary widely in Europe, countries of Finno-Ugrian origin show disproportionally higher suicide rates than the rest of Europe, suggesting a genetic cause^42^. Voracek (2006)^43^, tested this hypothesis in the U.S.A. using state-level self-reported ancestry from census data. The study found support for this hypothesis using historical data from 1913-1924 and 1928-1932, but not from 1990-1994. Taken together, these findings encourage an analysis with higher geospatial resolution of the sample (e.g. ZIP or county level) and a genetic characterization of the individuals ancestries, since self-reporting may be inaccurate. We also found that living in a ZIP code with higher proportion of *Native Hawaiian and other pacific islander females between ages 45 to 54* was associated with suicide attempts. These results are consistent with Ji and colleagues (2020)^44^ finding, who showed that Asian or Pacific Islander ancestry was a risk factor for suicide in healthcare professionals.

Seasonality of suicide attempts is also a well-known phenomenon, and climate has been suggested to potentially be the main cause of this pattern^45^. However, results have varied depending on the methods and location used^46,47^. Several studies have shown that solar radiation, precipitation, and temperature are associated with suicide risk^4–6^. In our results we observed that precipitation in April, solar radiation in July *(which represents solar radiation in* June, through September) and wind speed in July (which represents wind speed in May through September) were protective, whereas wind speed in April (which represents wind speed for the rest of the year) and maximum temperature (with lower temperatures) in December (which represents average temperature from October through April, minimum temperature from November through March, and maximum temperature from October through March) were associated with increased risk among patients 60 years of age or above. In patients under 60 years of age, precipitation in January (which represents precipitation December through February) and *water vapor in May* (which represents water vapor from April through October and minimum temperature in May) were protective, whereas *solar radiation in July (which represents solar radiation in* June through September), and *wind speed throughout the year* were associated with increased risk. Furthermore, while climatic variation exists even within zip code-level resolution, climatic features exhibit the closest overlap between individual-level and zip code-level features.

Another goal of the present work was to explore decreased restraint and access to means for suicide attempts using the number of alcohol and firearms vendors per 10,000 residents as proxies. Importantly, regardless of age, we observed that living in areas with more alcohol and firearm vendors was associated with increased risk for attempting suicide (albeit less so than several of the climatic and socio-demographic features identified as important). These results provide additional evidence regarding the importance of access to means as one of several risk factors for suicide, especially in the U.S., where a larger proportion of suicides are committed using firearms in comparison to other high income countries where only 5% use them^13,48,49^. Our findings also confirm the importance of including alcohol abuse in models of suicide risk^50,51^.

### Study Limitations and Future Directions

The present study had a number of limitations that should be considered when interpreting these findings. First, we utilized a cross-sectional design, and a lifetime suicide attempts variable. Thus, additional work is still needed to examine the degree to which the features identified in the present study might be predictive of future suicide attempts. Second, our findings regarding alcohol and firearms vendors should be interpreted cautiously, as there are several other variables (**Supplementary figure 7**) that could also potentially explain their association with suicide attempts which should be considered in future work. Third, while the present study provides clear evidence that zip code-level geospatial variables can predict suicide attempt risk better than random chance, the degree to which these variables might predict suicide attempts above and beyond well-validated individual-level risk factors (e.g., psychiatric diagnosis) remains unknown. While such work is beyond the scope of the present study, future work is still needed to ascertain if and how geospatial variables might interact with individual-level data to predict suicide risk.

## Conclusion

In this study we demonstrated the use of X-AI to explore the impact of more than 1,700 demographic and climatic features on suicide attempt risk with high geospatial resolution. This research provides additional evidence for the role of several demographic and climatic features in suicide attempts and demonstrates the utility of using geospatial features within an X-AI framework to improve suicide risk prediction.

## Supporting information

MVP Suicide Exemplar Workgroup

## Data Availability

All data produced in the present study are available upon reasonable request to the authors

## Acknowledgements

This research is based on data from the Million Veteran Program, Office of Research and Development (ORD), Veterans Health Administration (VHA), and was supported by award #I01CX001729 from the Clinical Science Research and Development (CSR&D) Service of VHA ORD. This publication does not represent the views of the Department of Veteran Affairs or the United States Government. We also thank and acknowledge MVP and the MVP Suicide Exemplar Workgroup for their contributions to this manuscript. This work was also supported in part by the joint U.S. Department of Veterans Affairs and US Department of Energy MVP CHAMPION program. This manuscript has been co-authored by UT-Battelle, LLC under contract no. DE-AC05-00OR22725 with the U.S. Department of Energy. The United States Government retains and the publisher, by accepting the article for publication, acknowledges that the United States Government retains a nonexclusive, paid-up, irrevocable, world-wide license to publish or reproduce the published form of this manuscript, or allow others to do so, for United States Government purposes. The Department of Energy will provide public access to these results of federally sponsored research in accordance with the DOE Public Access Plan (http://energy.gov/downloads/doe-public-access-plan, last accessed September 16, 2020).

