## Supplementary figures for "Using Explainable-AI to Find Geospatial Environmental and Sociodemographic Predictors of Suicide Attempts"

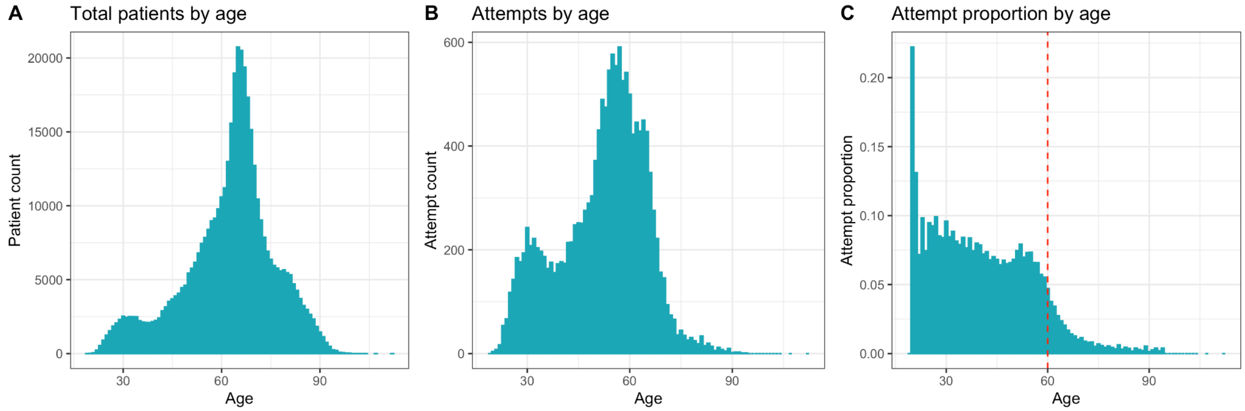


**Supplementary Figure 1. Distribution of suicide attempts by age. A.** Total patient count by age, **B.** Total suicide attempt count by age and **C.** Age-specific suicide rate. The red dashed line shows a cutoff to highlight the decreased proportion of suicide attempts in individuals equal to/above 60 years of age.

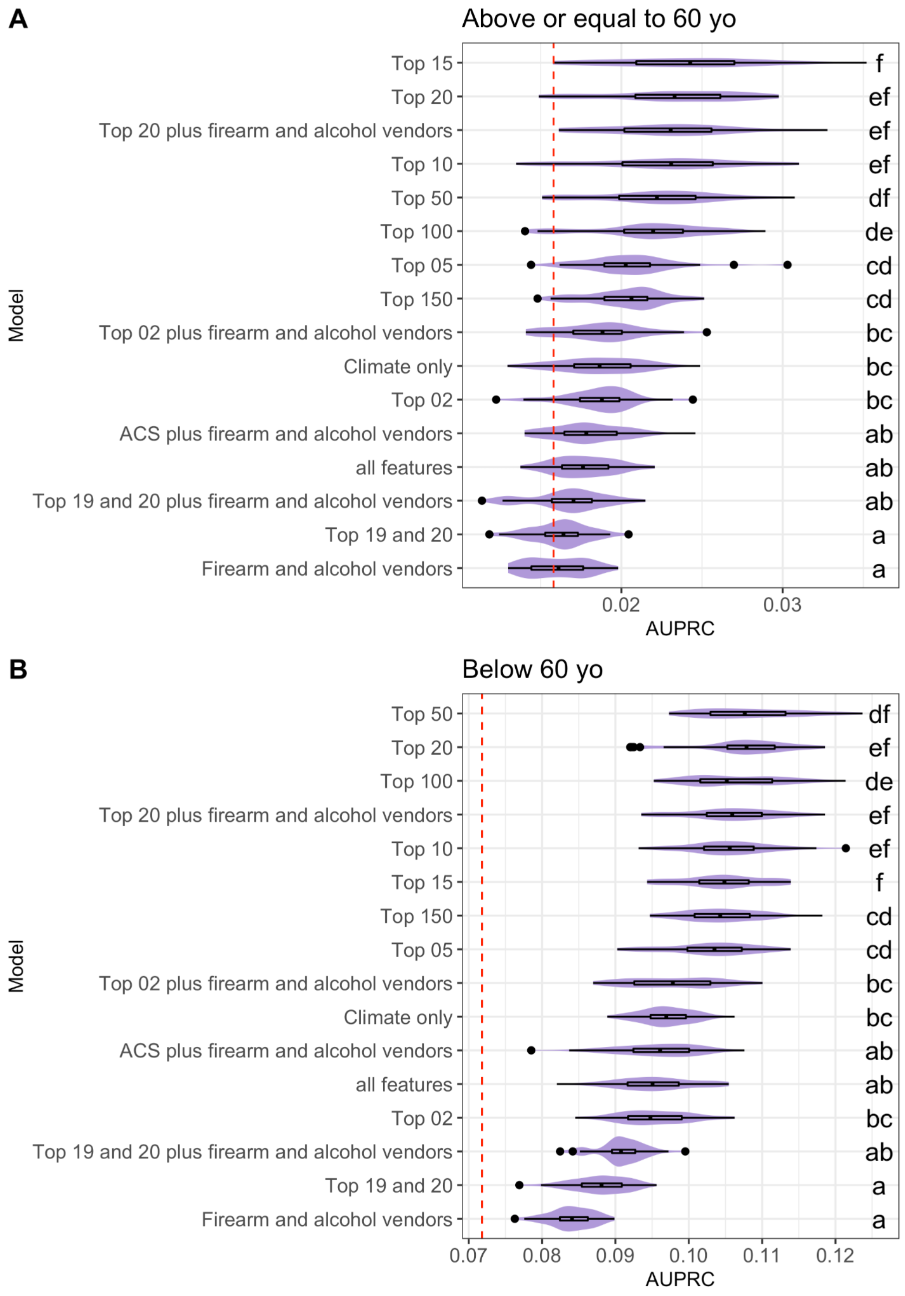


**Supplementary Figure 2.** Distribution of the area under the precision recall curves (AUPRC) for suicide attempt predictive capacity using environmental features. **A.** AUPRC values distributions for equal to/above 60 years of age for iRF cross validation models. **B.** AUPRC values for below 60 years of age. The red dashed line is the area under the curve of a base model (random chance of correct classification without iRF).

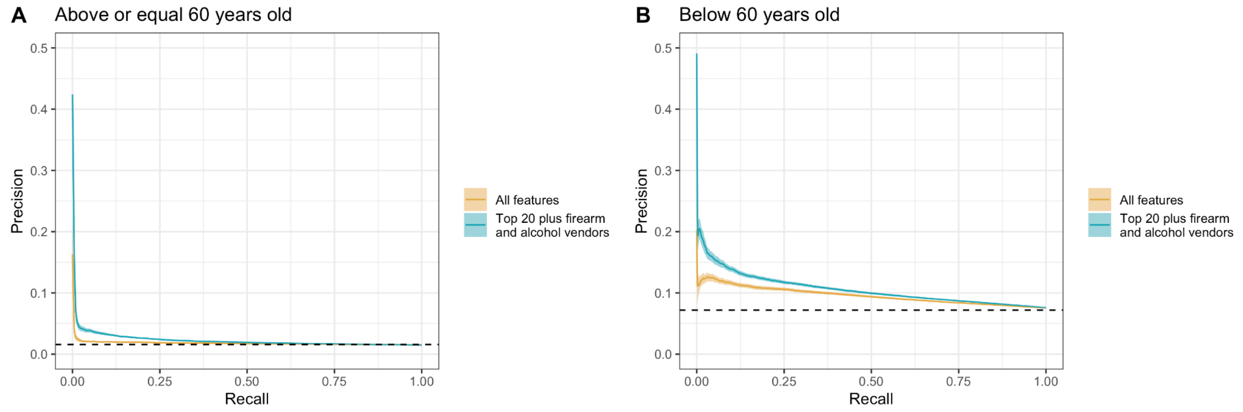


**Supplementary Figure 3**. Precision-recall curves for an iRF model using all features (red line) vs using only the top 20 features plus firearm and alcohol vendors per 10,000 residents (blue line). Each line represents the average of 50 runs.

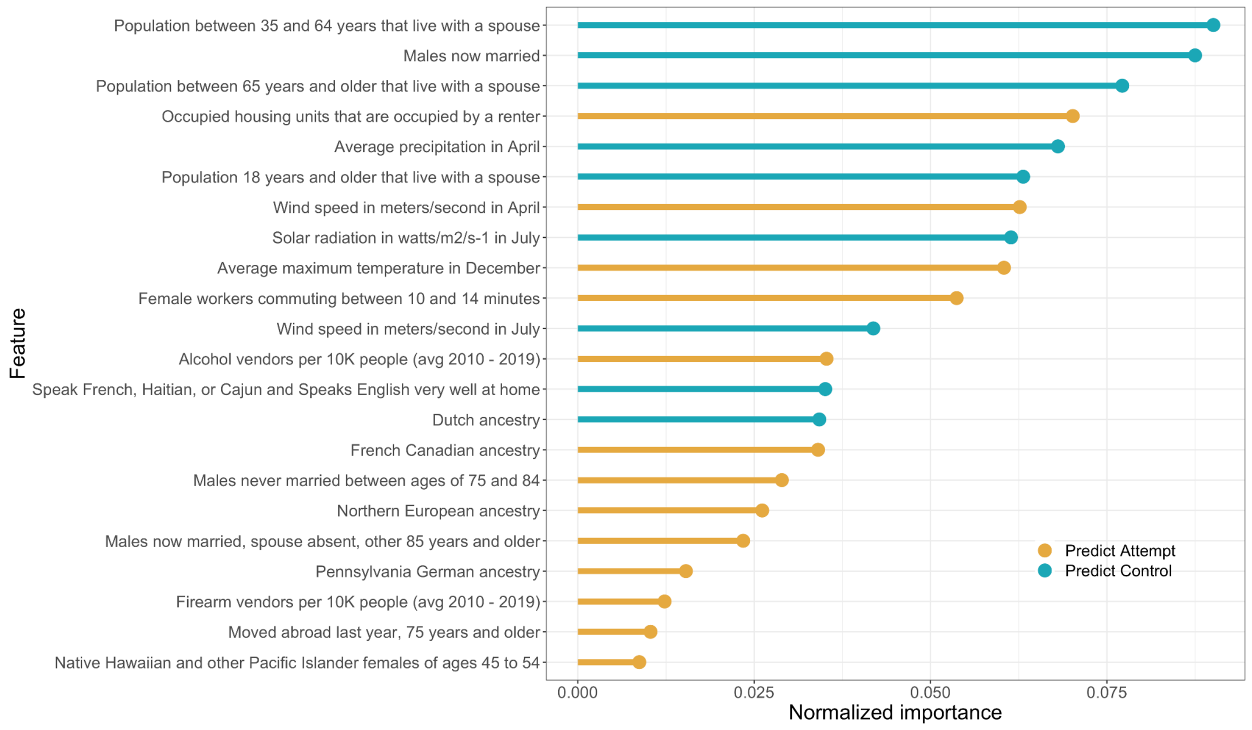


**Supplementary Figure 4.** Normalized importance of features in a suicide attempt iRF model for patients equal to/above 60 years of age, using the top 20 socio-demographic and climatic features plus the number of alcohol and firearm vendors per 10,000 people. Yellow: zip code-level features predicting subjects with a history of suicide attempt, Teal: features predictive of controls (no history of suicide attempt).

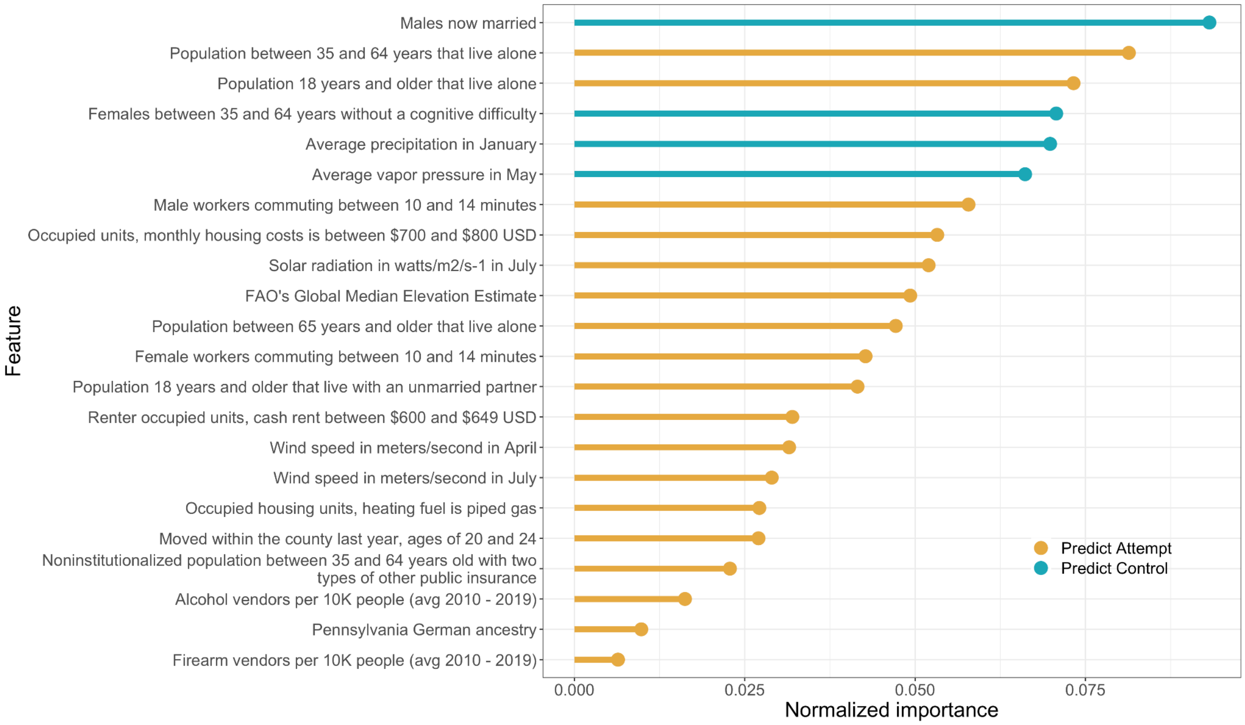


**Supplementary Figure 5.** Normalized importance of features in a predictive suicide attempt iRF model for patients under 60 years of age, using the top 20 socio-demographic and climatic features plus the number of alcohol and firearm vendors per 10,000 people. Yellow: zip code-level features predicting subjects with a history of suicide attempt, Teal: features predictive of controls (no history of suicide attempt).

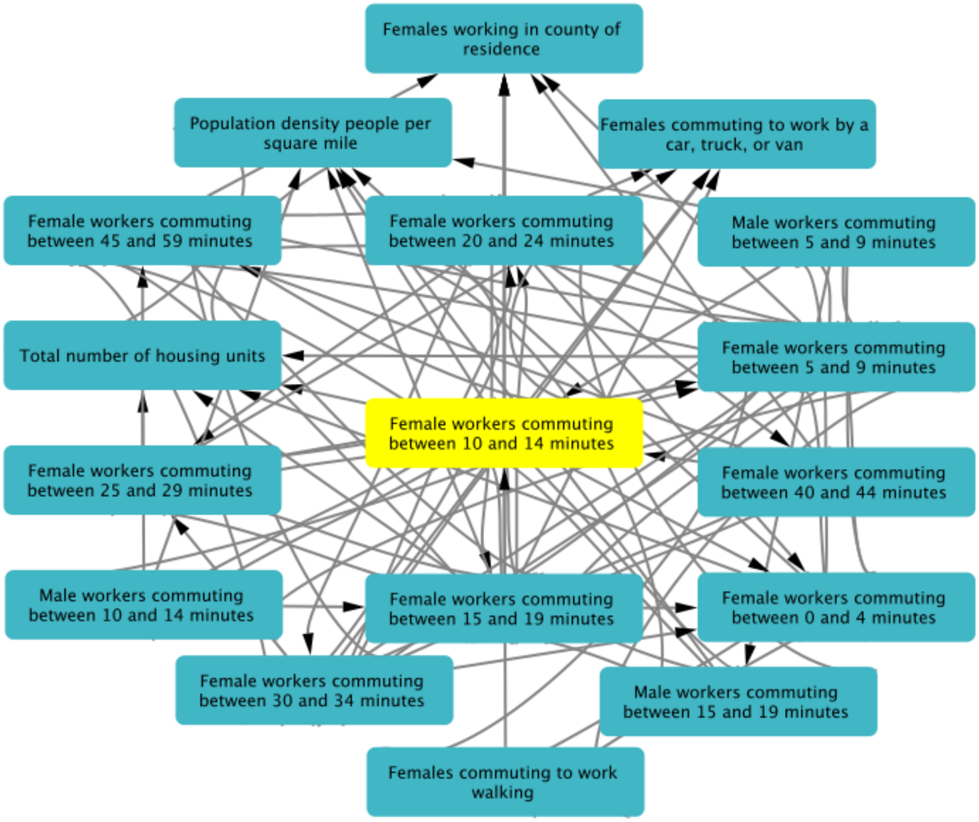


**Supplementary Figure 6**. iRF-LOOP subnetwork of the first neighbors for the proportion of females commuting 10 - 14 minutes to work.

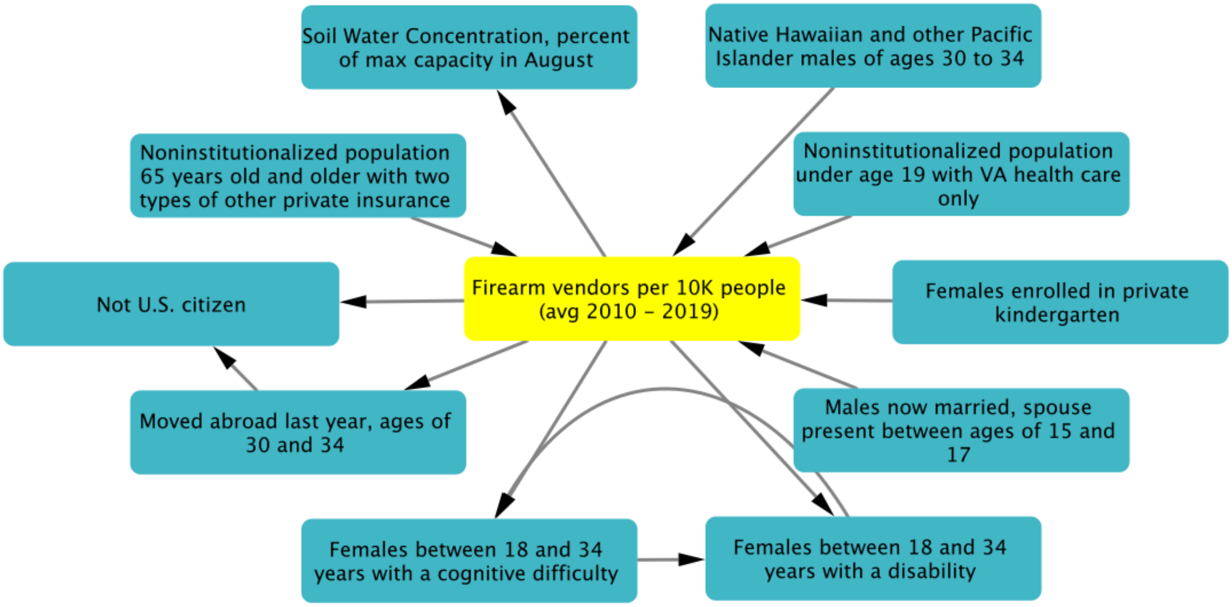


**Supplementary Figure 7.** iRF-LOOP subnetwork of the first neighbors for the number of firearm vendors per 10,000 residents.
