## Supplementary material for "Using Explainable-AI to Find Geospatial Environmental and Sociodemographic Predictors of Suicide Attempts": MVP Suicide Exemplar Workgroup

### VA Million Veteran Program (MVP) Acknowledgements


MVP Executive Committee

- Co-Chair: J. Michael Gaziano, M.D., M.P.H.
  VA Boston Healthcare System, 150 S. Huntington Avenue, Boston, MA 02130
- Co-Chair: Sumitra Muralidhar, Ph.D.
  US Department of Veterans Affairs, 810 Vermont Avenue NW, Washington, DC 20420
- Rachel Ramoni, D.M.D., Sc.D., Chief VA Research and Development Officer
  US Department of Veterans Affairs, 810 Vermont Avenue NW, Washington, DC 20420
- Jean C. Beckham, Ph.D.
  Durham VA Medical Center, 508 Fulton Street, Durham, NC 27705
- Kyong-Mi Chang, M.D.
  Philadelphia VA Medical Center, 3900 Woodland Avenue, Philadelphia, PA 19104
- Christopher J. O'Donnell, M.D., M.P.H.
  VA Boston Healthcare System, 150 S. Huntington Avenue, Boston, MA 02130
- Philip S. Tsao, Ph.D.
  VA Palo Alto Health Care System, 3801 Miranda Avenue, Palo Alto, CA 94304
- James Breeling, M.D., Ex-Officio
  US Department of Veterans Affairs, 810 Vermont Avenue NW, Washington, DC 20420
- Grant Huang, Ph.D., Ex-Officio
  US Department of Veterans Affairs, 810 Vermont Avenue NW, Washington, DC 20420
- Juan P. Casas, M.D., Ph.D., Ex-Officio
  VA Boston Healthcare System, 150 S. Huntington Avenue, Boston, MA 02130

MVP Program Office

- Sumitra Muralidhar, Ph.D.
  US Department of Veterans Affairs, 810 Vermont Avenue NW, Washington, DC 20420
- Jennifer Moser, Ph.D.
  US Department of Veterans Affairs, 810 Vermont Avenue NW, Washington, DC 20420

MVP Recruitment/Enrollment

- Recruitment/Enrollment Director/Deputy Director, Boston – Stacey B. Whitbourne, Ph.D.; Jessica V. Brewer, M.P.H.
  VA Boston Healthcare System, 150 S. Huntington Avenue, Boston, MA 02130
- MVP Coordinating Centers
  - Clinical Epidemiology Research Center (CERC), West Haven – Mihaela Aslan, Ph.D.
    West Haven VA Medical Center, 950 Campbell Avenue, West Haven, CT 06516
  - Cooperative Studies Program Clinical Research Pharmacy Coordinating Center, Albuquerque – Todd Connor, Pharm.D.; Dean P. Argyres, B.S., M.S.
    New Mexico VA Health Care System, 1501 San Pedro Drive SE, Albuquerque, NM 87108
  - Genomics Coordinating Center, Palo Alto – Philip S. Tsao, Ph.D.
    VA Palo Alto Health Care System, 3801 Miranda Avenue, Palo Alto, CA 94304
  - MVP Boston Coordinating Center, Boston - J. Michael Gaziano, M.D., M.P.H.

VA Boston Healthcare System, 150 S. Huntington Avenue, Boston, MA 02130
o MVP Information Center, Canandaigua – Brady Stephens, M.S.
Canandaigua VA Medical Center, 400 Fort Hill Avenue, Canandaigua, NY 14424

- VA Central Biorepository, Boston – Mary T. Brophy M.D., M.P.H.; Donald E. Humphries, Ph.D.; Luis E. Selva, Ph.D.
  VA Boston Healthcare System, 150 S. Huntington Avenue, Boston, MA 02130
- MVP Informatics, Boston – Nhan Do, M.D.; Shahpoor (Alex) Shayan, M.S.
  VA Boston Healthcare System, 150 S. Huntington Avenue, Boston, MA 02130
- MVP Data Operations/Analytics, Boston – Kelly Cho, M.P.H., Ph.D.
  VA Boston Healthcare System, 150 S. Huntington Avenue, Boston, MA 02130
- Director of Regulatory Affairs – Lori Churby, B.S.
  VA Palo Alto Health Care System, 3801 Miranda Avenue, Palo Alto, CA 94304

MVP Science

- Science Operations – Christopher J. O'Donnell, M.D., M.P.H.

VA Boston Healthcare System, 150 S. Huntington Avenue, Boston, MA 02130

- Genomics Core – Christopher J. O'Donnell, M.D., M.P.H.; Saiju Pyarajan Ph.D.
  VA Boston Healthcare System, 150 S. Huntington Avenue, Boston, MA 02130
  Philip S. Tsao, Ph.D.
  VA Palo Alto Health Care System, 3801 Miranda Avenue, Palo Alto, CA 94304
- Data Core – Kelly Cho, M.P.H, Ph.D.
  VA Boston Healthcare System, 150 S. Huntington Avenue, Boston, MA 02130
- VA Informatics and Computing Infrastructure (VINCI) – Scott L. DuVall, Ph.D.
  VA Salt Lake City Health Care System, 500 Foothill Drive, Salt Lake City, UT 84148
- Data and Computational Sciences – Saiju Pyarajan, Ph.D.
  VA Boston Healthcare System, 150 S. Huntington Avenue, Boston, MA 02130
- Statistical Genetics – Elizabeth Hauser, Ph.D.
  Durham VA Medical Center, 508 Fulton Street, Durham, NC 27705
  Yan Sun, Ph.D.
  Atlanta VA Medical Center, 1670 Clairmont Road, Decatur, GA 30033
  Hongyu Zhao, Ph.D.
  West Haven VA Medical Center, 950 Campbell Avenue, West Haven, CT 06516

Current MVP Local Site Investigators

- Atlanta VA Medical Center (Peter Wilson, M.D.)
  1670 Clairmont Road, Decatur, GA 30033
- Bay Pines VA Healthcare System (Rachel McArdle, Ph.D.)
  10,000 Bay Pines Blvd Bay Pines, FL 33744
- Birmingham VA Medical Center (Louis Dellitalia, M.D.)
  700 S. 19th Street, Birmingham AL 35233
- Central Western Massachusetts Healthcare System (Kristin Mattocks, Ph.D., M.P.H.)
  421 North Main Street, Leeds, MA 01053
- Cincinnati VA Medical Center (John Harley, M.D., Ph.D.)
  3200 Vine Street, Cincinnati, OH 45220

- Clement J. Zablocki VA Medical Center (Jeffrey Whittle, M.D., M.P.H.)
  5000 West National Avenue, Milwaukee, WI 53295
- VA Northeast Ohio Healthcare System (Frank Jacono, M.D.)
  10701 East Boulevard, Cleveland, OH 44106
- Durham VA Medical Center (Jean Beckham, Ph.D.)
  508 Fulton Street, Durham, NC 27705
- Edith Nourse Rogers Memorial Veterans Hospital (John Wells., Ph.D.)
  200 Springs Road, Bedford, MA 01730
- Edward Hines, Jr. VA Medical Center (Salvador Gutierrez, M.D.)
  5000 South 5th Avenue, Hines, IL 60141
- Veterans Health Care System of the Ozarks (Gretchen Gibson, D.D.S., M.P.H.)
  1100 North College Avenue, Fayetteville, AR 72703
- Fargo VA Health Care System (Kimberly Hammer, Ph.D.)
  2101 N. Elm, Fargo, ND 58102
- VA Health Care Upstate New York (Laurence Kaminsky, Ph.D.)
  113 Holland Avenue, Albany, NY 12208
- New Mexico VA Health Care System (Gerardo Villareal, M.D.)
  1501 San Pedro Drive, S.E. Albuquerque, NM 87108
- VA Boston Healthcare System (Scott Kinlay, M.B.B.S., Ph.D.)
  150 S. Huntington Avenue, Boston, MA 02130
- VA Western New York Healthcare System (Junzhe Xu, M.D.)
  3495 Bailey Avenue, Buffalo, NY 14215-1199
- Ralph H. Johnson VA Medical Center (Mark Hamner, M.D.)
  109 Bee Street, Mental Health Research, Charleston, SC 29401
- Columbia VA Health Care System (Roy Mathew, M.D.)
  6439 Garners Ferry Road, Columbia, SC 29209
- VA North Texas Health Care System (Sujata Bhushan, M.D.)
  4500 S. Lancaster Road, Dallas, TX 75216
- Hampton VA Medical Center (Pran Iruvanti, D.O., Ph.D.)
  100 Emancipation Drive, Hampton, VA 23667
- Richmond VA Medical Center (Michael Godschalk, M.D.)
  1201 Broad Rock Blvd., Richmond, VA 23249
- Iowa City VA Health Care System (Zuhair Ballas, M.D.)
  601 Highway 6 West, Iowa City, IA 52246-2208
- Eastern Oklahoma VA Health Care System (Douglas Ivins, M.D.)
  1011 Honor Heights Drive, Muskogee, OK 74401
- James A. Haley Veterans' Hospital (Stephen Mastorides, M.D.)
  13000 Bruce B. Downs Blvd, Tampa, FL 33612
- James H. Quillen VA Medical Center (Jonathan Moorman, M.D., Ph.D.)
  Corner of Lamont & Veterans Way, Mountain Home, TN 37684
- John D. Dingell VA Medical Center (Saib Gappy, M.D.)
  4646 John R Street, Detroit, MI 48201
- Louisville VA Medical Center (Jon Klein, M.D., Ph.D.)
  800 Zorn Avenue, Louisville, KY 40206

- Manchester VA Medical Center (Nora Ratcliffe, M.D.)
  718 Smyth Road, Manchester, NH 03104
- Miami VA Health Care System (Hermes Florez, M.D., Ph.D.)
  1201 NW 16th Street, 11 GRC, Miami FL 33125
- Michael E. DeBakey VA Medical Center (Olaoluwa Okusaga, M.D.)
  2002 Holcombe Blvd, Houston, TX 77030
- Minneapolis VA Health Care System (Maureen Murdoch, M.D., M.P.H.)
  One Veterans Drive, Minneapolis, MN 55417
- N. FL/S. GA Veterans Health System (Peruvemba Sriram, M.D.)
  1601 SW Archer Road, Gainesville, FL 32608
- Northport VA Medical Center (Shing Shing Yeh, Ph.D., M.D.)
  79 Middleville Road, Northport, NY 11768
- Overton Brooks VA Medical Center (Neeraj Tandon, M.D.)
  510 East Stoner Ave, Shreveport, LA 71101
- Philadelphia VA Medical Center (Darshana Jhala, M.D.)
  3900 Woodland Avenue, Philadelphia, PA 19104
- Phoenix VA Health Care System (Samuel Aguayo, M.D.)
  650 E. Indian School Road, Phoenix, AZ 85012
- Portland VA Medical Center (David Cohen, M.D.)
  3710 SW U.S. Veterans Hospital Road, Portland, OR 97239
- Providence VA Medical Center (Satish Sharma, M.D.)
  830 Chalkstone Avenue, Providence, RI 02908
- Richard Roudebush VA Medical Center (Suthat Liangpunsakul, M.D., M.P.H.)
  1481 West 10th Street, Indianapolis, IN 46202
- Salem VA Medical Center (Kris Ann Oursler, M.D.)
  1970 Roanoke Blvd, Salem, VA 24153
- San Francisco VA Health Care System (Mary Whooley, M.D.)
  4150 Clement Street, San Francisco, CA 94121
- South Texas Veterans Health Care System (Sunil Ahuja, M.D.)
  7400 Merton Minter Boulevard, San Antonio, TX 78229
- Southeast Louisiana Veterans Health Care System (Joseph Constans, Ph.D.)
  2400 Canal Street, New Orleans, LA 70119
- Southern Arizona VA Health Care System (Paul Meyer, M.D., Ph.D.)
  3601 S 6th Avenue, Tucson, AZ 85723
- Sioux Falls VA Health Care System (Jennifer Greco, M.D.)
  2501 W 22nd Street, Sioux Falls, SD 57105
- St. Louis VA Health Care System (Michael Rauchman, M.D.)
  915 North Grand Blvd, St. Louis, MO 63106
- Syracuse VA Medical Center (Richard Servatius, Ph.D.)
  800 Irving Avenue, Syracuse, NY 13210
- VA Eastern Kansas Health Care System (Melinda Gaddy, Ph.D.)
  4101 S 4th Street Trafficway, Leavenworth, KS 66048
- VA Greater Los Angeles Health Care System (Agnes Wallbom, M.D., M.S.)
  11301 Wilshire Blvd, Los Angeles, CA 90073

- VA Long Beach Healthcare System (Timothy Morgan, M.D.)
  5901 East 7th Street Long Beach, CA 90822
- VA Maine Healthcare System (Todd Stapley, D.O.)
  1 VA Center, Augusta, ME 04330
- VA New York Harbor Healthcare System (Scott Sherman, M.D., M.P.H.)
  423 East 23rd Street, New York, NY 10010
- VA Pacific Islands Health Care System (George Ross, M.D.)
  459 Patterson Rd, Honolulu, HI 96819
- VA Palo Alto Health Care System (Philip Tsao, Ph.D.)
  3801 Miranda Avenue, Palo Alto, CA 94304-1290
- VA Pittsburgh Health Care System (Patrick Strollo, Jr., M.D.)
  University Drive, Pittsburgh, PA 15240
- VA Puget Sound Health Care System (Edward Boyko, M.D.)
  1660 S. Columbian Way, Seattle, WA 98108-1597
- VA Salt Lake City Health Care System (Laurence Meyer, M.D., Ph.D.)
  500 Foothill Drive, Salt Lake City, UT 84148
- VA San Diego Healthcare System (Samir Gupta, M.D., M.S.C.S.)
  3350 La Jolla Village Drive, San Diego, CA 92161
- VA Sierra Nevada Health Care System (Mostaqul Huq, Pharm.D., Ph.D.)
  975 Kirman Avenue, Reno, NV 89502
- VA Southern Nevada Healthcare System (Joseph Fayad, M.D.)
  6900 North Pecos Road, North Las Vegas, NV 89086
- VA Tennessee Valley Healthcare System (Adriana Hung, M.D., M.P.H.)
  1310 24th Avenue, South Nashville, TN 37212
- Washington DC VA Medical Center (Jack Lichy, M.D., Ph.D.)
  50 Irving St, Washington, D. C. 20422
- W.G. (Bill) Hefner VA Medical Center (Robin Hurley, M.D.)
  1601 Brenner Ave, Salisbury, NC 28144
- White River Junction VA Medical Center (Brooks Robey, M.D.)
  163 Veterans Drive, White River Junction, VT 05009
- William S. Middleton Memorial Veterans Hospital (Robert Striker, M.D., Ph.D.)
  2500 Overlook Terrace, Madison, WI 53705

#### MVP Suicide Exemplar Workgroup Acknowledgements

The MVP Suicide Exemplar Workgroup for this publication includes Khushbu Agarwal, Allison E. Ashley-Koch, Mihaela Aslan, Jean C. Beckham, Edmond Begoli, Tanmoy Bhattacharya, Ben Brown, Patrick S. Calhoun, Mikaela Cashman McDevitt, Kei-Hoi Cheung, Sutanay Choudhury, Ashley M. Cliff, Judith D. Cohn, Silvia Crivelli, Leticia Cuellar-Hengartner, Haedi E. Deangelis, Michelle F. Dennis, Sayera Dhaubhadel, Patrick D. Finley, Kumkum Ganguly, Michael R. Garvin, Joel E. Gelernter, Lauren P. Hair, Phillip D. Harvey, Elizabeth R. Hauser, Michael A. Hauser, Nick W. Hengartner, Daniel A. Jacobson, Piet C. Jones, David Kainer, Alan D. Kaplan, Ira R. Katz, Rachel L. Kember, Nathan A. Kimbrel, Angela C. Kirby, John C. Ko, Beauty Kolade, John Lagergren, Matthew Lane, Daniel F. Levey, Drew Levin, Jennifer H. Lindquist, Xianlian Liu, Ravi

K. Madduri, Carrie Manore, Susana B. Martins, John F. McCarthy, Benjamin H. McMahon, J. Izaak Miller, Destinee Morrow, David W. Oslin, Mirko Pavicic-Venegas, John P. Pestian, Saiju Pyarajan, Xue J. Qin, Nallakkandi Rajeevan, Christine M. Ramsey, Ruy Ribeiro, Jonathon Romero, Alex Rodriguez, Daniel Santel, Noah Schaefferkoetter, Yunling Shi, Murray B. Stein, Kyle A. Sullivan, Ning Sun, Suzanne R. Tamang, Alice Townsend, Jodie A. Trafton, Angelica Walker, Xiange Wang, Victoria Wangia-Anderson, Renji Yang, Shinjae Yoo, Hong-Jun Yoon, Rafael Zamora-Resendiz, and Hongyu Zhao.

#### International Suicide Genetics Consortium (ISGC) Acknowledgements

Niamh Mullins[1, 2], Jooeun Kang [3], Adrian I. Campos [4, 5], Jonathan R. I. Coleman [6, 7], Alexis C. Edwards [8], Hanga Galfalvy [9, 10], Daniel F. Levey [11, 12], Adriana Lori [13], Andrey Shabalin [14], Anna Starnawska [15, 16, 17, 18], Mei-Hsin Su [19], Hunna J. Watson [20, 21, 22], Mark Adams [23], Swapnil Awasthi [24], Michael Gandal [25], Jonathan D. Hafferty [23], Akitoyo Hishimoto [26], Minsoo Kim [25], Satoshi Okazaki [27], Ikuo Otsuka [10, 27], Stephan Ripke [24, 28, 29], Erin B. Ware [30, 31], Andrew W. Bergen [32, 33], Wade H. Berrettini [34,] Martin Bohus [35], Harry Brandt [36], Xiao Chang [37], Wei J. Chen [19, 38, 39], Hsi-Chung Chen [39], Steven Crawford [36], Scott Crow [40], Emily DiBlasi [14], Philibert Duriez [41, 42], Fernando Fernández-Aranda [43], Manfred M. Fichter [44, 45], Steven Gallinger [46], Stephen J. Glatt [47], Philip Gorwood [41, 42], Yiran Guo [37], Hakon Hakonarson [37, 48], Katherine A. Halmi [49], Hai-Gwo Hwu [50], Sonia Jain [51], Stéphane Jamain [52], Susana Jiménez-Murcia [43], Craig Johnson [53], Allan S. Kaplan [54, 55, 56], Walter H. Kaye [57], Pamela K. Keel [58], James L. Kennedy [54, 55, 56], Kelly L. Klump [59], Robert D. Levitan [54, 55, 56], Dong Li [37], Shih-Cheng Liao [39], Klaus Lieb [60], Lisa Lilenfeld [61], Chih-Min Liu [39], Pierre J. Magistretti [62, 63], Christian R. Marshall [64], James E. Mitchell [65], Eric T. Monson [14], Richard M. Myers [66], Dalila Pinto [1, 2], Abigail Powers [13], Nicolas Ramoz [42], Stefan Roepke [67], Alessandro Rotondo [68], Vsevolod Rozanov [69, 70], Stephen W. Scherer [71], Christian Schmahl [35], Marcus Sokolowski [72], Michael Strober [73, 74], Laura M. Thornton [22], Janet Treasure [75, 76], Ming T. Tsuang [77], Maria C La Via [22], Stephanie H Witt [78], D. Blake Woodside [55, 56, 79, 80], Zeynep Yilmaz [22, 81, 82], Lea Zillich [78], Rolf Adolfsson [83], Ingrid Agartz [84, 85, 86], Tracy M. Air [87], Martin Alda [88, 89], Lars Alfredsson [90, 91], Ole A. Andreassen [92, 93], Adebayo Anjorin [94], Vivek Appadurai [95, 96], María Soler Artigas [97, 98, 99, 100], Sandra Van der Auwera [101], M. Helena Azevedo [102], Nicholas Bass [103], Claiton H.D. Bau [104, 105], Bernhard T. Baune [106, 107], Frank Bellivier [108, 109, 110, 111], Klaus Berger [112], Joanna M. Biernacka [113], Tim B. Bigdeli [114, 115], Elisabeth B. Binder [13, 116], Michael Boehnke [117], Marco P. Boks [118], Rosa Bosch [97, 98, 119], David L. Braff [120], Richard Bryant [121], Monika Budde [122], Enda M. Byrne [123, 124], Wiepke Cahn [125], Miguel Casas [97, 98, 100, 119], Enrique Castelao [126], Jorge A. Cervilla [127], Boris Chaumette [128, 129, 130], Sven Cichon [131, 132, 133, 134], Aiden Corvin [135], Nicholas Craddock [136], David Craig [137], Franziska Degenhardt [134], Srdjan Djurovic [138, 139], Howard J. Edenberg [140, 141], Ayman H. Fanous [114, 115], Jerome C. Foo [142], Andreas J. Forstner [131, 134, 143], Mark Frye [144], Janice M Fullerton [145, 146], Justine M Gatt [121, 145], Pablo V. Gejman [147, 148], Ina Giegling [149, 150], Hans J. Grabe [101], Melissa J. Green [145, 151], Eugenio H. Grevet [152, 153], Maria Grigoroiu-Serbanescu [154], Blanca Gutierrez [155], Jose Guzman-Parra [156], Steven P. Hamilton [157], Marian L. Hamshere [136], Annette Hartmann [149], Joanna Hauser [158], Stefanie Heilmann-Heimbach [134], Per Hoffmann [132, 133, 134], Marcus Ising [159], Ian Jones [136], Lisa A. Jones [160], Lina Jonsson [161], René S. Kahn [2, 162], John R. Kelsoe [120, 163], Kenneth S. Kendler [115], Stefan Kloiber [54, 159, 164], Karestan C. Koenen [165, 166, 167], Manolis Kogevinas [168], Bettina Konte [149], Marie-Odile Krebs [128, 129, 130], Mikael Landén [161, 169], Jacob

Lawrence [170], Marion Leboyer [108, 171, 172], Phil H Lee [28, 29, 173], Douglas F. Levinson [174], Calwing Liao [175, 176], Jolanta Lissowska [177], Susanne Lucae [159], Fermin Mayoral [156], Susan L McElroy [178], Patrick McGrath [179], Peter McGuffin [7], Andrew McQuillin [103], Sarah E Medland [180], Divya Mehta [181, 182], Ingrid Melle [92, 183], Yuri Milaneschi [184], Philip B. Mitchell [151], Esther Molina [185], Gunnar Morken [186, 187], Preben Bo Mortensen [16, 81, 96, 188], Bertram Müller-Myhsok [116, 189, 190], Caroline Nievergelt [120], Vishwajit Nimgaonkar [191], Markus M. Nöthen [134], Michael C. O'Donovan [136], Roel A. Ophoff [192, 193], Michael J. Owen [136], Carlos Pato [194, 195], Michele T. Pato [195], Brenda W.J.H. Penninx [184], Jonathan Pimm [103], Giorgio Pistis [126], James B. Potash [196], Robert A. Power [7, 197, 198], Martin Preisig [126], Digby Quested [199], Josep Antoni Ramos-Quiroga [97, 98, 100, 119], Andreas Reif [200], Marta Ribasés [97, 98, 99, 100], Vanesa Richarte [97, 98, 119], Marcella Rietschel [201], Margarita Rivera [7, 202], Andrea Roberts [203], Gloria Roberts [151], Guy A. Rouleau [176, 204], Diego L. Rovaris [205], Dan Rujescu [149], Cristina Sánchez-Mora [97, 98, 99, 100], Alan R. Sanders [147, 148], Peter R. Schofield [145, 146], Thomas G. Schulze [122, 142, 206, 207, 208], Laura J. Scott [117], Alessandro Serretti [209], Jianxin Shi [210], Stanley I. Shyn [211], Lea Sirignano [142], Pamela Sklar [1, 2, 212], Olav B. Smeland [92, 93], Jordan W. Smoller [28, 167, 213], Edmund J.S. Sonuga-Barke [214], Gianfranco Spalletta [215, 216], John S. Strauss [54, 164], Beata Świątkowska [217], Maciej Trzaskowski [123], Gustavo Turecki [218], Laura Vilar-Ribó [97, 100], John B. Vincent [219], Henry Völzke [220], James T.R. Walters [136], Cynthia Shannon Weickert [145, 151], Thomas W. Weickert [145, 151], Myrna M Weissman [221, 222], Leanne M. Williams [223], Naomi R. Wray [123, 182], Clement C. Zai [28, 164, 166, 224, 225, 226], Allison E. Ashley-Koch [227], Jean C. Beckham [228, 229], Elizabeth R. Hauser [227, 230], Michael A. Hauser [227], Nathan A. Kimbrel [228, 229], Jennifer H. Lindquist [231], Benjamin McMahon [232], David W. Oslin [233, 234], Xuejun Qin [227], Major Depressive Disorder Working Group of the Psychiatric Genomics Consortium, Bipolar Disorder Working Group of the Psychiatric Genomics Consortium, Eating Disorders Working Group of the Psychiatric Genomics Consortium, German Borderline Genomics Consortium, MVP Suicide Exemplar Workgroup, VA Million Veteran Program, Esben Agerbo [81, 188, 235], Anders D Børglum [15, 16, 17, 18], Gerome Breen [6, 7], Annette Erlangsen [18, 236, 237, 238], Tõnu Esko [239, 240], Joel Gelernter [11, 12], David M. Hougaard [235, 241], Ronald C. Kessler [242], Henry R. Kranzler [243, 244], Qingqin S. Li [245], Nicholas G. Martin [246], Andrew M. McIntosh [23], Ole Mors [235, 247], Merete Nordentoft [235, 248], Catherine M. Olsen [249], David Porteous [250], Robert J. Ursano [251], Danuta Wasserman [72], Thomas Werge [95, 235, 252, 253], David C. Whiteman [249], Cynthia M. Bulik [22, 169, 254], Hilary Coon [14, 255], Ditte Demontis [15, 16, 17, 18], Anna R. Docherty [8, 14], Po-Hsiu Kuo [19, 39], Cathryn M. Lewis [7, 256], J. John Mann [257], Miguel E. Rentería [4, 5], Daniel J. Smith [258], Eli A. Stahl [1, 2, 239], Murray B. Stein [259], Fabian Streit [78], Virginia Willour [260], Douglas M. Ruderfer [3, 261, 262]

**Affiliations**

[1] Department of Genetics and Genomic Sciences, Icahn School of Medicine at Mount Sinai, New York, NY, USA

[2] Department of Psychiatry, Icahn School of Medicine at Mount Sinai, New York, NY, USA

[3] Division of Genetic Medicine, Department of Medicine, Vanderbilt Genetics Institute, Vanderbilt University Medical Center, Nashville, TN, USA

[4] Department of Genetics and Computational Biology, QIMR Berghofer Medical Research Institute, Brisbane, QLD, Australia

[5] School of Biomedical Sciences, Faculty of Medicine, The University of Queensland, Brisbane, QLD, Australia

[6] National Institute for Health Research (NIHR) Maudsley Biomedical Research Centre at South London and Maudsley NHS Foundation Trust, King's College London, London, UK
[7] Social Genetic and Developmental Psychiatry Centre, King's College London, London, UK
[8] Department of Psychiatry, Virginia Commonwealth University, Richmond, VA, USA
[9] Department of Biostatistics, Columbia University, New York, NY, USA
[10] Department of Psychiatry, Columbia University, New York, NY, USA
[11] Department of Psychiatry, Veterans Affairs Connecticut Healthcare Center, West Haven, CT, USA
[12] Division of Human Genetics, Department of Psychiatry, Yale University School of Medicine, New Haven, CT, USA
[13] Department of Psychiatry and Behavioral Sciences, Emory University School of Medicine, Atlanta, GA, USA
[14] Department of Psychiatry, University of Utah School of Medicine, Salt Lake City, UT, USA
[15] Centre for Genomics and Personalized Medicine, CGPM, Aarhus University, Aarhus, Denmark
[16] Centre for Integrative Sequencing, iSEQ, Aarhus University, Aarhus, Denmark
[17] Department of Biomedicine, Aarhus University, Aarhus, Denmark
[18] The Lundbeck Foundation Initiative for Integrative Psychiatric Research, iPSYCH, Aarhus University, Aarhus, Denmark
[19] Institute of Epidemiology and Preventive Medicine, College of Public Health, National Taiwan University, Taipei, Taiwan
[20] School of Psychology, Curtin University, Perth, Western Australia, Australia
[21] Division of Paediatrics, The University of Western Australia, Perth, Western Australia, Australia
[22] Department of Psychiatry, University of North Carolina at Chapel Hill, Chapel Hill, NC, USA
[23] Division of Psychiatry, University of Edinburgh, Edinburgh, UK
[24] Department of Psychiatry and Psychotherapy, Charité - Universitätsmedizin Berlin, Berlin, Germany
[25] Department of Psychiatry and Biobehavioral Science, Semel Institute, David Geffen School of Medicine, University of California, Los Angeles, Los Angeles, CA, USA
[26] Department of Psychiatry, Yokohama City University Graduate School of Medicine, Yokohama, Japan
[27] Department of Psychiatry, Kobe University Graduate School of Medicine, Kobe, Japan
[28] Stanley Center for Psychiatric Research, Broad Institute, Cambridge, MA, USA
[29] Analytical and Translational Genetics Unit, Massachusetts General Hospital, Boston, MA, USA
[30] Population Studies Center, Institute for Social Research, University of Michigan, Ann Arbor, MI, USA
[31] Survey Research Center, Institute for Social Research, University of Michigan, Ann Arbor, MI, USA
[32] BioRealm, LLC, Walnut, CA, USA
[33] Oregon Research Institute, Eugene, OR, USA
[34] Department of Psychiatry, Center for Neurobiology and Behavior, Perelman School of Medicine at the University of Pennsylvania, Philadelphia, PA, USA
[35] Department of Psychosomatic Medicine and Psychotherapy, Central Institute of Mental Health, Medical Faculty Mannheim, University of Heidelberg, Mannheim, Germany
[36] The Center for Eating Disorders at Sheppard Pratt, Baltimore, MD, USA

[37] Center for Applied Genomics, Children's Hospital of Philadelphia, Philadelphia, PA, USA
[38] Center for Neuropsychiatric Research, National Health Research Institutes, Miaoli County, Taiwan
[39] Department of Psychiatry, National Taiwan University Hospital, Taipei, Taiwan
[40] Department of Psychiatry, University of Minnesota, Minneapolis, MN, USA
[41] Hôpital Sainte Anne, GHU Paris Psychiatrie et Neurosciences, Paris, France
[42] Institute of Psychiatry and Neuroscience of Paris (IPNP), INSERM U1266, Université de Paris, Paris, France
[43] Department of Psychiatry, University Hospital Bellvitge-IDIBELL and CIBEROBN, Barcelona, Spain
[44] Department of Psychiatry and Psychotherapy, Ludwig-Maximilians-University (LMU), Munich, Germany
[45] Schön Klinik Roseneck affiliated with the Medical Faculty of the University of Munich (LMU), Munich, Germany
[46] Department of Surgery, Faculty of Medicine, University of Toronto, Toronto, ON, Canada
[47] Department of Psychiatry and Behavioral Sciences, SUNY Upstate Medical University, Syracuse, NY, USA
[48] The Perelman School of Medicine, University of Pennsylvania, Philadelphia, PA, USA
[49] Department of Psychiatry, Weill Cornell Medical College, New York, NY, USA
[50] Department of Psychiatry, National Taiwan University Hospital and College of Medicine, Taipei, Taiwan
[51] Biostatistics Research Center, Herbert Wertheim School of Public Health and Human Longevity Science, University of California San Diego, La Jolla, CA, USA
[52] Inserm U955, Institut Mondor de recherches Biomédicales, Laboratoire, Neuro-Psychiatrie Translationnelle, and Fédération Hospitalo-Universitaire de Précision Médecine en Addictologie et Psychiatrie (FHU ADAPT), University Paris-Est-Créteil, Créteil, France
[53] Eating Recovery Center, Denver, CO, USA
[54] Centre for Addiction and Mental Health, Toronto, ON, Canada
[55] Department of Psychiatry, University of Toronto, Toronto, ON, Canada
[56] Institute of Medical Science, University of Toronto, Toronto, ON, Canada
[57] Department of Psychiatry, University of California San Diego, San Diego, CA, USA
[58] Department of Psychology, Florida State University, Tallahassee, FL, USA
[59] Department of Psychology, Michigan State University, Lansing, MI, USA
[60] Department of Psychiatry and Psychotherapy, University Medical Center, Mainz, Germany
[61] Department of Clinical Psychology, The Chicago School of Professional Psychology, Washington, DC, USA
[62] BESE Division, King Abdullah University of Science and Technology, Thuwal, Saudi Arabia
[63] Department of Psychiatry, University of Lausanne-University Hospital of Lausanne (UNIL-CHUV), Lausanne, Switzerland
[64] Department of Paediatric Laboratory Medicine, The Hospital for Sick Children, Toronto, ON, Canada
[65] Department of Psychiatry and Behavioral Science, University of North Dakota School of Medicine and Health Sciences, Fargo, ND, USA
[66] HudsonAlpha Institute for Biotechnology, Huntsville, AL, USA

[67] Department of Psychiatry, Charité - Universitätsmedizin Berlin, Corporate Member of Freie Universität Berlin, Humboldt-Universität zu Berlin, Berlin Institute of Health, Campus Benjamin Franklin, Berlin, Germany
[68] Department of Psychiatry, Neurobiology, Pharmacology, and Biotechnologies, University of Pisa, Pisa, Italy
[69] Department of Psychology, Saint-Petersburg State University, Saint-Petersburg, Russian Federation
[70] Department of Borderline Disorders and Psychotherapy, V.M. Bekhterev National Medical Research Center for Psychiatry and Neurology, Saint-Petersburg, Russian Federation
[71] Department of Genetics and Genomic Biology, The Hospital for Sick Children, Toronto, ON, Canada
[72] National Centre for Suicide Research and Prevention of Mental Ill-Health (NASP), LIME, Karolinska Institutet, Stockholm, Sweden
[73] David Geffen School of Medicine, University of California Los Angeles, Los Angeles, CA, USA
[74] Department of Psychiatry and Biobehavioral Science, Semel Institute for Neuroscience and Human Behavior, University of California Los Angeles, Los Angeles, CA, USA
[75] Institute of Psychiatry, Psychology and Neuroscience, Department of Psychological Medicine, King's College London, London, UK
[76] National Institute for Health Research Biomedical Research Centre, King's College London and South London and Maudsley National Health Service Foundation Trust, London, UK
[77] Center for Behavioral Genomics, Department of Psychiatry, University of California, San Diego, San Diego, CA, USA
[78] Department of Genetic Epidemiology in Psychiatry, Central Institute of Mental Health, Medical Faculty Mannheim, University of Heidelberg, Mannheim, Germany
[79] Centre for Mental Health, University Health Network, Toronto, ON, Canada
[80] Program for Eating Disorders, University Health Network, Toronto, ON, Canada
[81] National Centre for Register-Based Research, Aarhus University, Aarhus, Denmark
[82] Department of Genetics, University of North Carolina at Chapel Hill, Chapel Hill, NC, USA
[83] Department of Clinical Sciences, Psychiatry, Umeå University Medical Faculty, Umeå, Sweden
[84] Department of Psychiatric Research, Diakonhjemmet Hospital, Oslo, Norway
[85] Department of Clinical Neuroscience, Centre for Psychiatry Research, Karolinska Institutet, Stockholm, Sweden
[86] NORMENT, Institute of Clinical Medicine, University of Oslo, Oslo, Norway
[87] Discipline of Psychiatry, University of Adelaide, Adelaide, SA, Australia
[88] Department of Psychiatry, Dalhousie University, Halifax, NS, Canada
[89] National Institute of Mental Health, Klecany, CZ
[90] Department of Clinical Neuroscience, Karolinska Institutet, Stockholm, Sweden
[91] Inst of Environmental Medicine, Karolinska Institutet, Stockholm, Sweden
[92] Division of Mental Health and Addiction, Oslo University Hospital, Oslo, Norway
[93] NORMENT, University of Oslo, Oslo, Norway
[94] Psychiatry, Berkshire Healthcare NHS Foundation Trust, Bracknell, UK
[95] Institute of Biological Psychiatry, Copenhagen Mental Health Services, Copenhagen University Hospital, Copenhagen, Denmark

[96] The Lundbeck Foundation Initiative for Integrative Psychiatric Research, iPSYCH, Copenhagen, Denmark
[97] Department of Psychiatry, Hospital Universitari Vall d'Hebron, Barcelona, Spain
[98] Biomedical Network Research Centre on Mental Health (CIBERSAM), Instituto de Salud Carlos III, Madrid, Spain
[99] Department of Genetics, Microbiology & Statistics, University of Barcelona, Barcelona, Spain
[100] Psychiatric Genetics Unit, Group of Psychiatry, Mental Health and Addiction, Vall d'Hebron Research Institute (VHIR), Universitat Autònoma de Barcelona, Barcelona, Spain
[101] Department of Psychiatry and Psychotherapy, University Medicine Greifswald, Greifswald, Mecklenburg-Vorpommern, Germany
[102] Department of Psychiatry, University of Coimbra, Coimbra, Portugal
[103] Division of Psychiatry, University College London, London, UK
[104] Laboratory of Developmental Psychiatry, Hospital de Clínicas de Porto Alegre, Porto Alegre, RS, Brazil
[105] Department of Genetics, Universidade Federal do Rio Grande do Sul, Porto Alegre, RS, Brazil
[106] Department of Psychiatry, Melbourne Medical School, University of Melbourne, Melbourne, Australia
[107] Department of Psychiatry, University of Münster, Münster, Germany
[108] Department of Psychiatry and Addiction Medicine, Assistance Publique - Hôpitaux de Paris, Paris, France
[109] Paris Bipolar and TRD Expert Centres, FondaMental Foundation, Paris, France
[110] UMR-S1144 Team 1: Biomarkers of relapse and therapeutic response in addiction and mood disorders, INSERM, Paris, France
[111] Psychiatry, Université Paris Diderot, Paris, France
[112] Institute of Epidemiology and Social Medicine, University of Münster, Münster, Nordrhein-Westfalen, Germany
[113] Health Sciences Research, Mayo Clinic, Rochester, MN, USA
[114] Department of Psychiatry and Behavioral Sciences, State University of New York Downstate Medical Center, New York, NY, USA
[115] Department of Psychiatry, Virginia Commonwealth University, Richmond, VA, USA
[116] Department of Translational Research in Psychiatry, Max Planck Institute of Psychiatry, Munich, Germany
[117] Center for Statistical Genetics and Department of Biostatistics, University of Michigan, Ann Arbor, MI, USA
[118] Psychiatry, UMC Utrecht Hersencentrum, Utrecht, Netherlands
[119] Department of Psychiatry and Legal Medicine, Universitat Autònoma de Barcelona, Barcelona, Spain
[120] Department of Psychiatry, University of California San Diego, La Jolla, CA, USA
[121] School of Psychology, University of New South Wales, Sydney, NSW, Australia
[122] Institute of Psychiatric Phenomics and Genomics (IPPG), University Hospital, LMU Munich, Munich, Germany
[123] Institute for Molecular Bioscience, The University of Queensland, Brisbane, QLD, Australia
[124] Centre for Children's Health Research, The University of Queensland, Brisbane, QLD, Australia

[125] Department of Psychiatry, UMC Utrecht Hersencentrum Rudolf Magnus, Utrecht, Netherlands
[126] Department of Psychiatry, Lausanne University Hospital and University of Lausanne, Lausanne, Vaud, Switzerland
[127] Mental Health Unit, Department of Psychiatry, Faculty of Medicine, Granada University Hospital Complex, University of Granada, Granada, Spain
[128] Institut de Psychiatrie, CNRS GDR 3557, Paris, France
[129] Department of Evaluation, Prevention and Therapeutic innovation, GHU Paris Psychiatrie et Neurosciences, Paris, France
[130] Team Pathophysiology of psychiatric diseases, Université de Paris, Institute of Psychiatry and Neuroscience of Paris (IPNP), INSERM U1266, Paris, France
[131] Institute of Neuroscience and Medicine (INM-1), Research Centre Jülich, Jülich, Germany
[132] Institute of Medical Genetics and Pathology, University Hospital Basel, Basel, Switzerland
[133] Department of Biomedicine, University of Basel, Basel, Switzerland
[134] Institute of Human Genetics, University of Bonn, School of Medicine & University Hospital Bonn, Bonn, Germany
[135] Neuropsychiatric Genetics Research Group, Dept of Psychiatry and Trinity Translational Medicine Institute, Trinity College Dublin, Dublin, Ireland
[136] Medical Research Council Centre for Neuropsychiatric Genetics and Genomics, Division of Psychological Medicine and Clinical Neurosciences, Cardiff University, Cardiff, UK
[137] Department of Translational Genomics, University of Southern California, Pasadena, CA, USA
[138] Department of Medical Genetics, Oslo University Hospital, Oslo, Norway
[139] NORMENT, KG Jebsen Centre for Psychosis Research, Department of Clinical Science, University of Bergen, Bergen, Norway
[140] Department of Medical & Molecular Genetics, Indiana University, Indianapolis, IN, USA
[141] Biochemistry and Molecular Biology, Indiana University School of Medicine, Indianapolis, IN, USA
[142] Department of Genetic Epidemiology in Psychiatry, Central Institute of Mental Health, Medical Faculty Mannheim, Heidelberg University, Mannheim, Germany
[143] Centre for Human Genetics, University of Marburg, Marburg, Germany
[144] Department of Psychiatry & Psychology, Mayo Clinic, Rochester, MN, USA
[145] Neuroscience Research Australia, Sydney, NSW, Australia
[146] School of Medical Sciences, University of New South Wales, Sydney, NSW, Australia
[147] Department of Psychiatry and Behavioral Sciences, NorthShore University HealthSystem, Evanston, IL, USA
[148] Department of Psychiatry and Behavioral Neuroscience, University of Chicago, Chicago, IL, USA
[149] Department of Psychiatry, Psychotherapy and Psychosomatics, Martin-Luther-University Halle-Wittenberg, Halle (Saale), Germany
[150] Department of Psychiatry, University of Munich, Munich, Germany
[151] School of Psychiatry, University of New South Wales, Sydney, NSW, Australia
[152] ADHD Outpatient Program, Adult Division, Hospital de Clínicas de Porto Alegre, Porto Alegre, RS, Brazil
[153] Department of Psychiatry, Universidade Federal do Rio Grande do Sul, Porto Alegre, RS, Brazil

[154] Biometric Psychiatric Genetics Research Unit, Alexandru Obregia Clinical Psychiatric Hospital, Bucharest, Romania
[155] Department of Psychiatry, Faculty of Medicine and Biomedical Research Centre (CIBM), University of Granada, Granada, Spain
[156] Mental Health Department, University Regional Hospital. Biomedicine Institute (IBIMA), Málaga, Spain
[157] Psychiatry, Kaiser Permanente Northern California, San Francisco, CA, USA
[158] Department of Psychiatry, Laboratory of Psychiatric Genetics, Poznan University of Medical Sciences, Poznan, Poland
[159] Max Planck Institute of Psychiatry, Munich, Germany
[160] Department of Psychological Medicine, University of Worcester, Worcester, UK
[161] Department of Psychiatry and Neuroscience, University of Gothenburg, Gothenburg, Sweden
[162] Psychiatry, UMC Utrecht Hersencentrum Rudolf Magnus, Utrecht, Netherlands
[163] Institute for Genomic Medicine, University of California San Diego, La Jolla, CA, USA
[164] Department of Psychiatry, University of Toronto, Toronto, ON, Canada
[165] Stanley Center for Psychiatric Research, Broad Institute, Cambridge, MA, USA
[166] Department of Epidemiology, Harvard TH Chan School of Public Health, Boston, MA, USA
[167] Department of Psychiatry, Massachusetts General Hospital, Boston, MA, USA
[168] Center for Research in Environmental Epidemiology (CREAL), Barcelona, Spain
[169] Department of Medical Epidemiology and Biostatistics, Karolinska Institutet, Stockholm, Sweden
[170] Psychiatry, North East London NHS Foundation Trust, Ilford, UK
[171] INSERM, Paris, France
[172] Faculté de Médecine, Université Paris Est, Créteil, France
[173] Psychiatric and Neurodevelopmental Genetics Unit, Massachusetts General Hospital, Boston, MA, USA
[174] Psychiatry & Behavioral Sciences, Stanford University, Stanford, CA, USA
[175] Department of Human Genetics, McGill University, Montreal, QC, Canada
[176] Montreal Neurological Institute and Hospital, Montreal, QC, Canada
[177] Cancer Epidemiology and Prevention, M. Sklodowska-Curie Cancer Center and Institute of Oncology, Warsaw, Poland
[178] Research Institute, Lindner Center of HOPE, Mason, OH, USA
[179] Psychiatry, Columbia University College of Physicians and Surgeons, New York, NY, USA
[180] Genetics and Computational Biology, QIMR Berghofer Medical Research Institute, Brisbane, QLD, Australia
[181] School of Psychology and Counseling, Queensland University of Technology, Brisbane, QLD, Australia
[182] Queensland Brain Institute, The University of Queensland, Brisbane, QLD, Australia
[183] Division of Mental Health and Addiction, University of Oslo, Institute of Clinical Medicine, Oslo, Norway
[184] Department of Psychiatry, Amsterdam UMC, Vrije Universiteit and GGZ inGeest, Amsterdam, Netherlands
[185] Department of Nursing, Faculty of Health Sciences and Biomedical Research Centre (CIBM), University of Granada, Granada, Spain

[186] Mental Health, Faculty of Medicine and Health Sciences, Norwegian University of Science and Technology - NTNU, Trondheim, Norway
[187] Psychiatry, St Olavs University Hospital, Trondheim, Norway
[188] Centre for Integrated Register-based Research, Aarhus University, Aarhus, Denmark
[189] Munich Cluster for Systems Neurology (SyNergy), Munich, Germany
[190] University of Liverpool, Liverpool, UK
[191] Psychiatry and Human Genetics, University of Pittsburgh, Pittsburgh, PA, USA
[192] Psychiatry, Erasmus University Medical Center, Rotterdam, Netherlands
[193] Jane and Terry Semel Institute for Neuroscience and Human Behavior, Los Angeles, CA, USA
[194] College of Medicine Institute for Genomic Health, SUNY Downstate Medical Center College of Medicine, Brooklyn, NY, USA
[195] Institute for Genomic Health, SUNY Downstate Medical Center College of Medicine, Brooklyn, NY, USA
[196] Psychiatry, University of Iowa, Iowa City, IA, USA
[197] Genetics, BioMarin Pharmaceuticals, London, UK
[198] St Edmund Hall, University of Oxford, Oxford, UK
[199] Department of Psychiatry, University of Oxford, Oxford, UK
[200] Department of Psychiatry, Psychosomatic Medicine and Psychotherapy, University Hospital Frankfurt, Frankfurt, Germany
[201] Department of Genetic Epidemiology in Psychiatry, Central Institute of Mental Health, Medical Faculty Mannheim, Heidelberg University, Mannheim, Baden-Württemberg, Germany
[202] Department of Biochemistry and Molecular Biology II and Institute of Neurosciences, Biomedical Research Centre (CIBM), University of Granada, Granada, Spain
[203] Department of Environmental Health, Harvard TH Chan School of Public Health, Boston, MA, USA
[204] Department of Neurology and Neurosurgery, McGill University, Faculty of Medicine, Montreal, QC, Canada
[205] Department of Physiology and Biophysics, Instituto de Ciencias Biomedicas Universidade de Sao Paulo, São Paulo, SP, Brazil
[206] Department of Psychiatry and Behavioral Sciences, Johns Hopkins University School of Medicine, Baltimore, MD, USA
[207] Human Genetics Branch, Intramural Research Program, National Institute of Mental Health, Bethesda, MD, USA
[208] Department of Psychiatry and Psychotherapy, University Medical Center Göttingen, Göttingen, Germany
[209] Department of Biomedical and NeuroMotor Sciences, University of Bologna, Bologna, Italy
[210] Division of Cancer Epidemiology and Genetics, National Cancer Institute, Bethesda, MD, USA
[211] Behavioral Health Services, Kaiser Permanente Washington, Seattle, WA, USA
[212] Department of Neuroscience, Icahn School of Medicine at Mount Sinai, New York, NY, USA
[213] Psychiatric and Neurodevelopmental Genetics Unit (PNGU), Massachusetts General Hospital, Boston, MA, USA
[214] Institute of Psychology, Psychiatry & Neuroscience, King's College London, London, UK
[215] Menninger Department of Psychiatry and Behavioral Sciences, Baylor College of Medicine, Houston, Houston, TX, USA

[216] Laboratory of Neuropsychiatry, IRCCS Santa Lucia Foundation, Rome, Rome, Italy
[217] Department of Environmental Epidemiology, Nofer Institute of Occupational Medicine, Lodz, Poland
[218] Department of Psychiatry, McGill University, Montreal, QC, Canada
[219] Molecular Brain Science, Centre for Addiction and Mental Health, Toronto, ON, Canada
[220] Institute for Community Medicine, University Medicine Greifswald, Greifswald, Mecklenburg-Vorpommern, Germany
[221] Columbia University College of Physicians and Surgeons, New York, NY, USA
[222] Division of Translational Epidemiology, New York State Psychiatric Institute, New York, NY, USA
[223] Department of Psychiatry and Behavioral Sciences, Stanford University, Stanford, CA, USA
[224] Institute of Medical Science, University of Toronto, Toronto, ON, Canada
[225] Molecular Brain Science, Campbell Family Mental Health Research Institute, Centre for Addiction and Mental Health, Toronto, ON, Canada
[226] Laboratory Medicine and Pathobiology, University of Toronto, Toronto, ON, Canada
[227] Duke Molecular Physiology Institute, Duke University Medical Center, Durham, NC, USA
[228] VISN 6 Mid-Atlantic Mental Illness Research, Education, and Clinical Center, Durham Veterans Affairs Health Care System, Durham, NC, USA
[229] Department of Psychiatry and Behavioral Sciences, Duke University School of Medicine, Durham, NC, USA
[230] Cooperative Studies Program Epidemiology Center, Durham Veterans Affairs Health Care System, Durham, NC, USA
[231] VA Health Services Research and Development Center of Innovation to Accelerate Discovery and Practice Transformation, Durham Veterans Affairs Health Care System, Durham, NC, USA
[232] Theoretical Biology and Biophysics, Los Alamos National Laboratory, Los Alamos National Laboratory, Los Alamos, NM, USA
[233] VISN 4 Mental Illness Research, Education, and Clinical Center, Corporal Michael J. Crescenz VA Medical Center, Philadelphia, PA, USA
[234] Department of Psychiatry, Perelman School of Medicine, University of Pennsylvania, Philadelphia, PA, USA
[235] The Lundbeck Foundation Initiative for Integrative Psychiatric Research, iPSYCH, Aarhus, Denmark
[236] Center of Mental Health Research, Australian National University, Canberra, Australia
[237] Department of Mental Health, Johns Hopkins Bloomberg School of Public Health, Baltimore, MD, USA
[238] Danish Research Institute for Suicide Prevention, Mental Health Centre Copenhagen, Copenhagen, Denmark
[239] Program in Medical and Population Genetics, Broad Institute, Cambridge, MA, USA
[240] Estonian Genome Center, Institute of Genomics, University of Tartu, Tartu, Estonia
[241] Center for Neonatal Screening, Department for Congenital Disorders, Statens Serum Institut, Copenhagen, Denmark
[242] Department of Health Care Policy, Harvard Medical School, Boston, MA, USA
[243] Department of Psychiatry, University of Pennsylvania Perelman School of Medicine, Philadelphia, PA, USA

[244] VISN 4 MIRECC, Crescenz VAMC, Philadelphia, PA, USA
[245] Neuroscience, Janssen Research & Development, LLC, Titusville, NJ, USA
[246] Department of Genetics and Computational Biology, QIMR Berghofer Medical Research Institute, Herston, QLD, Australia
[247] Psychosis Research Unit, Aarhus University Hospital, Risskov, Aarhus, Denmark
[248] Mental Health Center Copenhagen, Copenhagen University Hospital, Copenhagen, Denmark
[249] Department of Population Health, QIMR Berghofer Medical Research Institute, Herston, QLD, Australia
[250] Institute for Genetics and Molecular Medicine, University of Edinburgh, Edinburgh, UK
[251] Department of Psychiatry, Uniformed University of the Health Sciences, Bethesda, MD, USA
[252] Department of Clinical Medicine, University of Copenhagen, Copenhagen, Denmark
[253] Lundbeck Foundation GeoGenetics Centre, GLOBE Institute, University of Copenhagen, Copenhagen, Denmark
[254] Department of Nutrition, University of North Carolina at Chapel Hill, Chapel Hill, NC, USA
[255] Biomedical Informatics, University of Utah School of Medicine, Salt Lake City, UT, USA
[256] Department of Medical & Molecular Genetics, King's College London, London, UK
[257] Departments of Psychiatry and Radiology, Columbia University, New York, NY, USA
[258] Institute of Health and Wellbeing, University of Glasgow, Glasgow, UK
[259] Department of Psychiatry and School of Public Health, University of California San Diego, La Jolla, CA, USA
[260] Department of Psychiatry, University of Iowa, Iowa City, IA, USA
[261] Department of Biomedical Informatics, Vanderbilt University Medical Center, Nashville, TN, USA
[262] Department of Psychiatry and Behavioral Sciences, Vanderbilt University Medical Center, Nashville, TN, USA
